# The Road to Recovery: Dose-Response Effects of CPAP on Real-World Driving Performance in Obstructive Sleep Apnoea

**DOI:** 10.64898/2026.01.20.26344436

**Authors:** Kieran O. Lee, Tristan A. Bekinschtein, Ian E. Smith

## Abstract

**Background:** Obstructive Sleep Apnoea (OSA) increases motor vehicle collision risk, and while Continuous Positive Airway Pressure (CPAP) considerably reduces this risk, it remains unclear how much CPAP use is required to fully normalise driving behaviour, and whether baseline (Pre-CPAP) driving style influences treatment response.

**Methods:** In a prospective observational study embedded within routine clinical care, real-world driving behaviour was recorded using a smartphone telematics application in patients undergoing diagnostic assessment for OSA and subsequent CPAP initiation. A total of 7,134 trips (>1,800 driving hours) were collected from 48 patients before and after CPAP initiation. Braking, acceleration, and turning events were quantified in terms of magnitude, duration, and frequency. Linear mixed-effects models assessed Pre- versus Post-CPAP differences, predictors of Post-CPAP driving, and dose–response effects of nightly CPAP adherence, adjusting for demographics, OSA severity, sleepiness, and trip characteristics.

**Results:** Following CPAP initiation, significant changes were observed in 6 of 9 driving metrics, indicating a shift toward sharper and less aggressive driving behaviour. Baseline (Pre-CPAP) driving strongly predicted Post-CPAP performance across all outcomes. Nightly CPAP adherence was associated with dose-dependent convergence toward control-like driving behaviour, with significant adherence × baseline interactions for braking and acceleration frequency measures. Drivers exhibiting both overly aggressive and overly cautious baseline styles showed behavioural normalisation with increasing CPAP use.

**Conclusions:** CPAP treatment is associated with measurable improvements in real-world driving behaviour, with recovery dependent on both baseline driving style and treatment adherence. These findings highlight substantial inter-individual heterogeneity in driving impairment and recovery in OSA. Smartphone-based telematics provide an objective, scalable method for assessing functional driving outcomes and may support personalised clinical advice and regulatory decision-making regarding driving fitness in patients with OSA.

## Introduction

Driving is a complex everyday activity that requires sustained attention, sensorimotor coordination, and rapid decision-making in dynamic environments. Impairments in these functions can substantially increase the risk of motor vehicle collisions, with important consequences for both individual safety and public health. Obstructive sleep apnoea (OSA) is a well-established risk factor for impaired driving, with untreated patients experiencing approximately a threefold increase in collision risk compared with the general population (Ellen et al., 2006; Tregear et al., 2009). However, although relative risk is elevated, the absolute risk for any individual with OSA remains low, creating persistent challenges for clinicians when advising patients about driving fitness.

In recognition of these challenges, several expert bodies have issued calls to action regarding OSA-related driving risk, most notably the European Respiratory Society (ERS) statement on sleep apnoea, sleepiness and driving risk (Bonsignore et al., 2021). The statement highlights the need for improved detection and quantification of driving risk, including clarification of the relative contributions of OSA severity and excessive daytime sleepiness (EDS). It emphasises the importance of assessing real-world driving outcomes and developing objective, standardised tools for clinical evaluation, and calls for investigation of how continuous positive airway pressure (CPAP) use relates to the normalisation of driving risk. Finally, the ERS advocates the use of emerging technologies to overcome current limitations in assessing everyday driving safety in individuals with OSA.

Evidence linking untreated OSA to an increased risk of motor vehicle collisions has been demonstrated across multiple methodological approaches, including database analyses (Findley et al., 1988; George C F P and Smiley A, 1999; Findley et al., 2000; Barbé et al., 2007; Mulgrew et al., 2008), self-reported incidents (Haraldsson, Carenfelt, Diderichsen, et al., 1990; Lloberes et al., 2000; Masa et al., 2000; Shiomi et al., 2002) and driving simulator studies (Findley et al., 1989; Haraldsson, Carenfelt, Laurell, et al., 1990; Findley et al., 1995; George et al., 1996; Juniper et al., 2000; Risser et al., 2000; Hack et al., 2001). Encouragingly, evidence from these same paradigms indicates that CPAP reduces crash and near-miss risk. A large database cohort (n=5,308) reported reductions in accidents (2.6% to 1.1%) and near misses (9.5% to 3.5%) after CPAP initiation (Coelho et al., 2024). Consistent with this, motor vehicle agency data show lower collision rates in treated OSA (Findley et al., 2000; George, 2001), and simulator studies demonstrate improved behavioural performance Post-treatment (Hack et al., 2001; Turkington et al., 2004; Orth et al., 2005).

However, despite meaningful average improvements, some patients continue to show residual impairment compared with controls (Vakulin et al., 2011), with substantial inter-individual variability in vulnerability to OSA-related driving impairment (Vakulin et al., 2022), where some individuals outperform controls (George et al., 1996). Although CPAP is highly effective in controlling obstructive events (Patil et al., 2019), many patients do not achieve recommended adherence thresholds (Kribbs et al., 1993; Sin et al., 2002; Lewis et al., 2004; NICE, 2021), meaning a substantial proportion labelled “on treatment” remain inadequately treated. Residual excessive daytime sleepiness also persists in some treated cohorts (Gasa et al., 2013; Dongol and Williams, 2016). In registry data, collision risk reductions are largely confined to adherent users: in a Swedish cohort (n=1,478), good compliance (>4 h/night) was associated with a 70% reduction in collisions, whereas poor compliance (<4 h/night) was associated with a 54% increase (Karimi et al., 2015). A French cohort similarly reported better driving safety with higher hours of CPAP use (Coelho et al., 2024).

More broadly, prior research has examined dose–response relationships between CPAP use and a range of clinical outcomes. In patients with severe OSA, the proportion achieving normal scores on subjective and objective sleepiness measures, including the Epworth Sleepiness Scale (ESS), Multiple Sleep Latency Test (MSLT), and Functional Outcomes of Sleep Questionnaire (FOSQ), increases with greater CPAP use, up to approximately seven hours per night (Weaver et al., 2007), a pattern replicated in more representative clinical cohorts (Antic et al., 2011). For cardiovascular outcomes, clinically meaningful improvements appear to occur at lower levels of CPAP use (Barbé et al., 2010; Gervès-Pinquié et al., 2022). However, a key limitation of this literature is the reliance on averaged nightly adherence, which obscures night-to-night variability and may fail to capture short-term effects relevant to next-day functioning. This is particularly pertinent for driving risk, as even a single missed night of CPAP has been associated with increased daytime sleepiness (Yang et al., 2006), EEG slowing, and impaired driving simulator performance (Filtness et al., 2012). As a result, the amount of CPAP use required to normalise real-world driving behaviour, and the impact of short-term adherence fluctuations, remain poorly defined.

Although informative, prior research on OSA-related driving impairment has several limitations. Crash statistics capture only rare events and provide little insight into more subtle but potentially informative changes in driving behaviour. Self-reported incidents may be biased, particularly in clinical contexts where disclosure could affect driving privileges. In addition, many studies have focused on highly selected populations, often restricted to males with severe OSA, limiting generalisability. Finally, despite their experimental utility, driving simulators struggle to replicate the complexity of real-world driving and have limited evidence for predicting actual collision risk (Bonsignore et al., 2021).

Advances in telematics now make it possible to collect real-world driving data unobtrusively using smartphone applications. These methods have been shown to capture meaningful behavioural variation across large populations, including applications in insurance risk modelling (Baecke and Bocca, 2017; Huang and Meng, 2019) and large-scale naturalistic driving datasets, where smoother driving behaviour has been associated with favourable outcomes such as higher passenger tipping rates (Chandar et al., 2019). Within clinical populations, smartphone-based driving metrics have been used to examine associations between driving behaviour and preclinical markers of Alzheimer’s disease (Babulal et al., 2021). In the context of OSA, a study of older adults reported that increasing OSA severity was associated with a higher rate of adverse driving events (Doherty et al., 2022). Although these findings are not readily generalisable to the broader OSA population, they illustrate the potential of telematics to assess real-world driving behaviour. Recently, we showed that driving data collected during OSA screening revealed more aggressive turning behaviour in patients with OSA and excessive daytime sleepiness compared with clinic controls (Lee et al., 2025). Together, these studies highlight the potential of smartphone-based telematics to provide objective, behaviourally grounded assessments of driving that may inform clinical evaluation of driving safety in OSA.

In this study, we collected real-world driving data using a smartphone application during clinical evaluation for suspected OSA and followed patients through diagnosis and initiation of CPAP therapy. We aimed to quantify changes in driving behaviour from Pre- to Post-CPAP and to examine dose–response relationships between nightly CPAP use and next-day driving performance.

## Methods

### Study Design and Participants

This was a prospective observational study measuring real-world driving behaviour before and after initiation of continuous positive airway pressure (CPAP) therapy for obstructive sleep apnoea (OSA). Participants were recruited from the Sleep Centre at Royal Papworth Hospital NHS Foundation Trust (Cambridge, UK) between August 2023 and August 2024. All patients referred for evaluation of suspected OSA during the recruitment period were invited to participate. Eligible participants were required to hold a valid driving licence, actively drive, be aged ≥18 years, and have no CPAP use within the preceding six months. Written informed consent was obtained remotely. See supplementary text a for full description of recruitment procedures.

Following enrolment, participants installed a smartphone application (“Insights”, Sentiance) to passively collect driving data. Participants were informed that study participation did not alter legal driving responsibilities and that all Driver and Vehicle Licensing Agency (DVLA) requirements remained applicable. Driving data were recorded from study entry through diagnostic assessment and, where applicable, for a minimum of two weeks following CPAP initiation. Participants with confirmed OSA who commenced CPAP formed the primary analysis cohort. Individuals without OSA or excessive daytime sleepiness were used to derive a control dataset.

Participants commencing CPAP were reminded to continue usual driving behaviour with the application active. Nightly CPAP adherence data were extracted from device memory cards after completion of the Post-CPAP observation period (figure 1).

**Figure 1:**
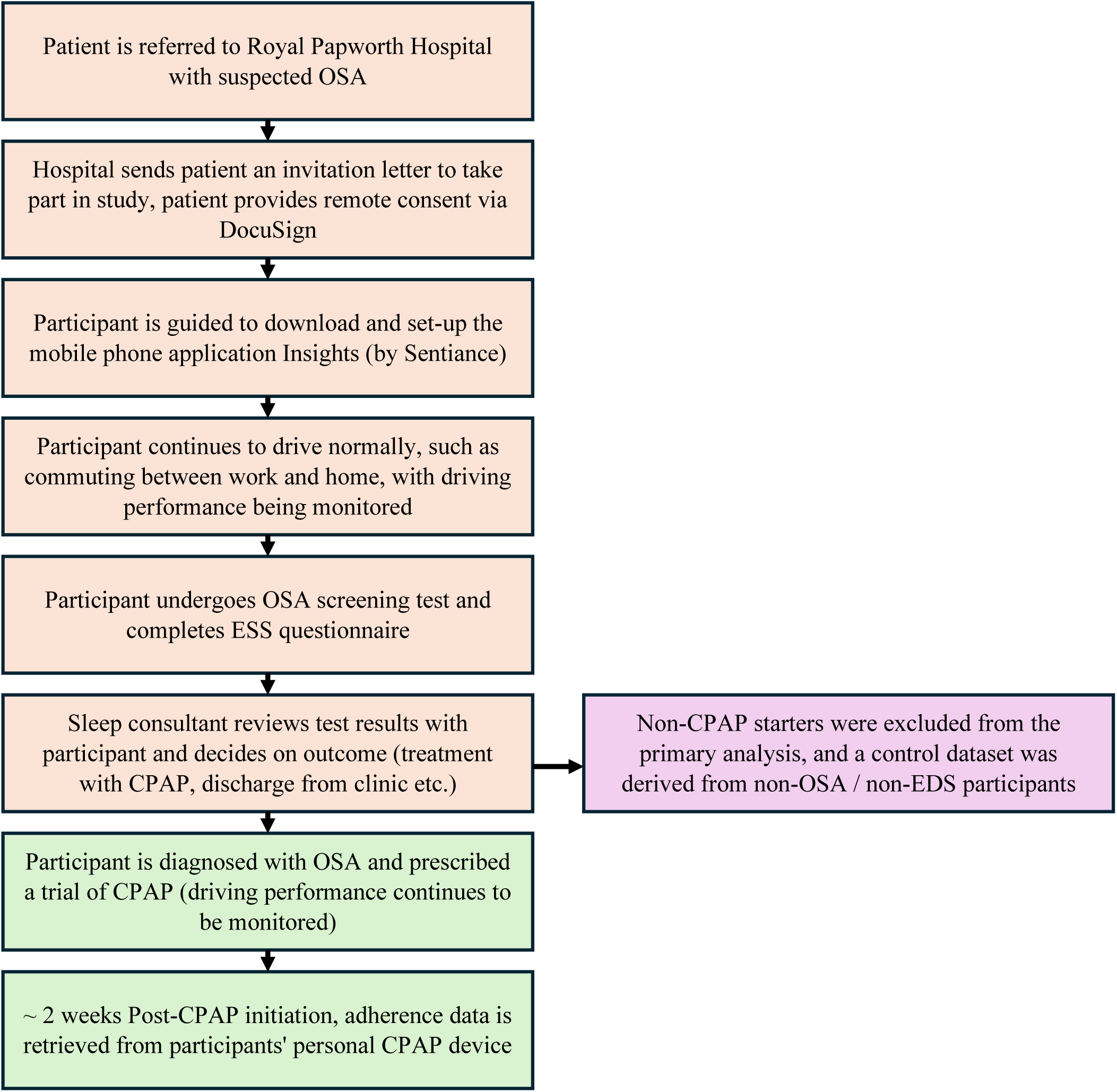
Flow-chart of study procedure.

Diagnostic and demographic data were extracted from the hospital electronic health record. OSA severity was assessed using the oxygen desaturation index (ODI) obtained from home pulse oximetry, which is the first-line diagnostic pathway at the study site, or the apnoea–hypopnoea index (AHI) where respiratory polygraphy or polysomnography was required. Subjective daytime sleepiness was assessed using the Epworth Sleepiness Scale (ESS), in line with routine clinical practice. The study received ethical approval from the UK Health Research Authority (30 May 2023; IRAS Project ID: 314762).

### Driving Performance Monitoring

The smartphone application captured motion data using the device’s built-in accelerometer and gyroscope, which were analysed to identify timestamped driving events corresponding to braking, acceleration, and turning. For each event, information on magnitude and duration was derived, and event frequency was calculated at the journey level (supplementary table a). Driving behaviours characterised by higher-magnitude events were interpreted as reflecting more aggressive driving.

### Data retention and statistical analysis

A total of 19,330 journeys were initially recorded from 98 participants, of whom 50 were diagnosed with OSA and prescribed CPAP therapy. After excluding participants who recorded no driving data and applying predefined trip-duration and Post-treatment exclusion criteria, the primary analysis dataset comprised 7,134 journeys (over 1,800 driving hours) from 48 participants. Of these, 3,559 trips were recorded Pre-CPAP and 3,575 Post-CPAP, with 33 participants contributing data in both conditions (supplementary figure a). Nightly CPAP adherence data were available for 31 participants and were matched to next-day driving, yielding a treatment adherence dataset of 2,232 trips (supplementary text b; supplementary table b).

A control dataset was derived from participants who screened negative for OSA (ODI <5) and excessive daytime sleepiness (Epworth Sleepiness Scale <11), comprising 1,353 journeys from 11 participants (supplementary text b).

Statistical analyses were performed using linear mixed-effects models, with participant included as a random effect to account for repeated measures. Fixed effects included trip duration and, where applicable, time into trip. Separate models were fitted for each driving metric. Effect sizes were derived from model estimates, and statistical significance was assessed using Bonferroni-adjusted p values (supplementary text c). Analyses were conducted in MATLAB (R2024a).

## Results

### Pre vs Post-CPAP treatment: Sharper but less aggressive driving

To assess changes in driving behaviour following CPAP initiation, linear mixed-effects models were fitted to 7,134 trips from 48 participants for each of the nine driving metrics. Compared with the Pre-CPAP period, Post-CPAP driving was associated with significant changes in six metrics: reduced brake duration (P < 0.0001, ES = −0.04), acceleration duration (P < 0.0001, ES = −0.03), significant acceleration frequency (P < 0.05, ES = −0.09), turn magnitude (P < 0.0001, ES = −0.03), turn duration (P < 0.0001, ES = −0.04), and significant turn frequency (P < 0.0001, ES = −0.13) (figure 2; supplementary table c). Together, these findings indicate a shift toward sharper (shorter-duration) but less aggressive (lower-magnitude and lower-frequency) driving behaviour following CPAP, with substantial inter-individual variability. Time into trip and trip duration were significant predictors across most outcomes, supporting their inclusion in the models.

**Figure 2:**
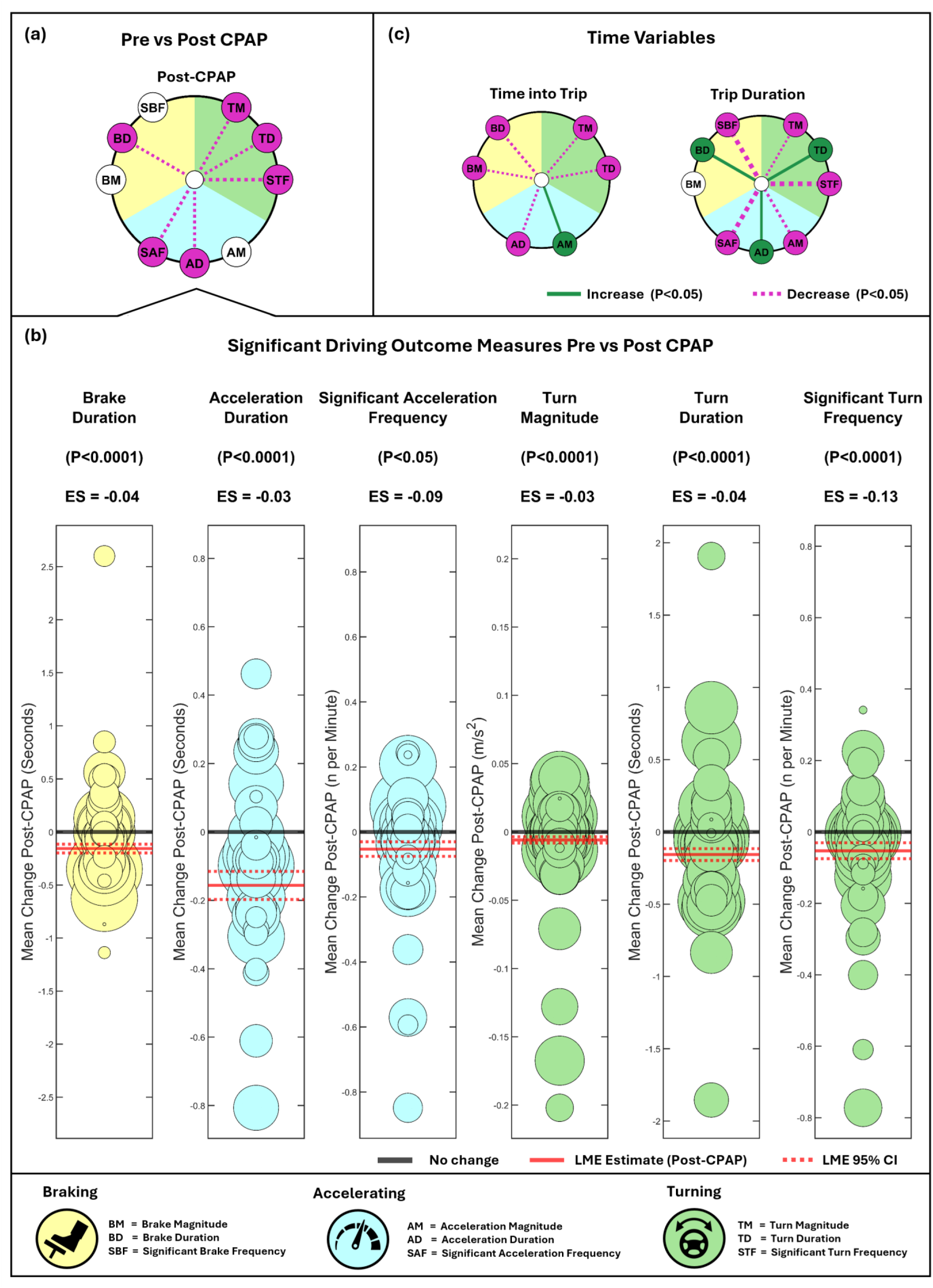
6 out of 9 driving performance measures showed differences Post-CPAP initiation. (a) Radial significance plots of linear mixed-effect (LME) model results testing Pre vs Post-CPAP. Each circle reflects independent fixed effects within models. Outer markers reflect driving outcome measures grouped by type: (Braking (yellow): BM = brake magnitude, BD = brake duration, SBF = significant brake frequency, Acceleration (blue): AM = acceleration magnitude, AD = acceleration duration, SAF = significant acceleration frequency and Turning (green): TM = turn magnitude, TD = turn duration, STF = significant turn frequency). Presence of a line indicates significance, green (increase), purple (decrease), width indicates effect size, cut at a maximum of 1.2 and a minimum of 0.05. (b) Plots show mean change in driving performance measures from Pre to Post-CPAP for each participant, across all measures significant at the group level. Size of marker reflects relative number of overall trips recorded by participant. Solid red line reflects the LME estimate Post-CPAP. Broken red line reflects the 95% confidence intervals of the LME estimate. (c) Radical significance plots for time variables (time into trip and trip duration), included in models.

### Post-CPAP with nightly compliance: Pre-CPAP driving strongly predicts Post-CPAP driving

To examine predictors of next-day driving behaviour, linear mixed-effects models were fitted including baseline (Pre-CPAP) driving performance, prior-night CPAP use, and time since CPAP initiation, adjusting for clinical, demographic, and trip-related covariates. Baseline driving performance was a significant predictor across all nine outcomes, with particularly strong effects for magnitude and significant frequency measures, indicating that Pre-treatment driving style was the strongest determinant of Post-CPAP behaviour among the variables considered.

Greater prior-night CPAP use was associated with changes in two outcomes, specifically reduced brake magnitude (P < 0.01, ES = −0.06) and reduced acceleration magnitude (P < 0.05, ES = −0.03), consistent with less aggressive driving with increasing nightly use. In contrast, longer treatment duration was associated with changes in three outcomes: reduced brake duration (P < 0.0001, ES = −0.07), reduced turn magnitude (P < 0.01, ES = −0.04), and reduced turn duration (P < 0.01, ES = −0.05) (figure 3; supplementary table d). These treatment-duration effects were directionally consistent with the Pre- versus Post-CPAP comparisons and suggest a shift toward sharper and less aggressive turning behaviour with increasing exposure to CPAP.

**Figure 3:**
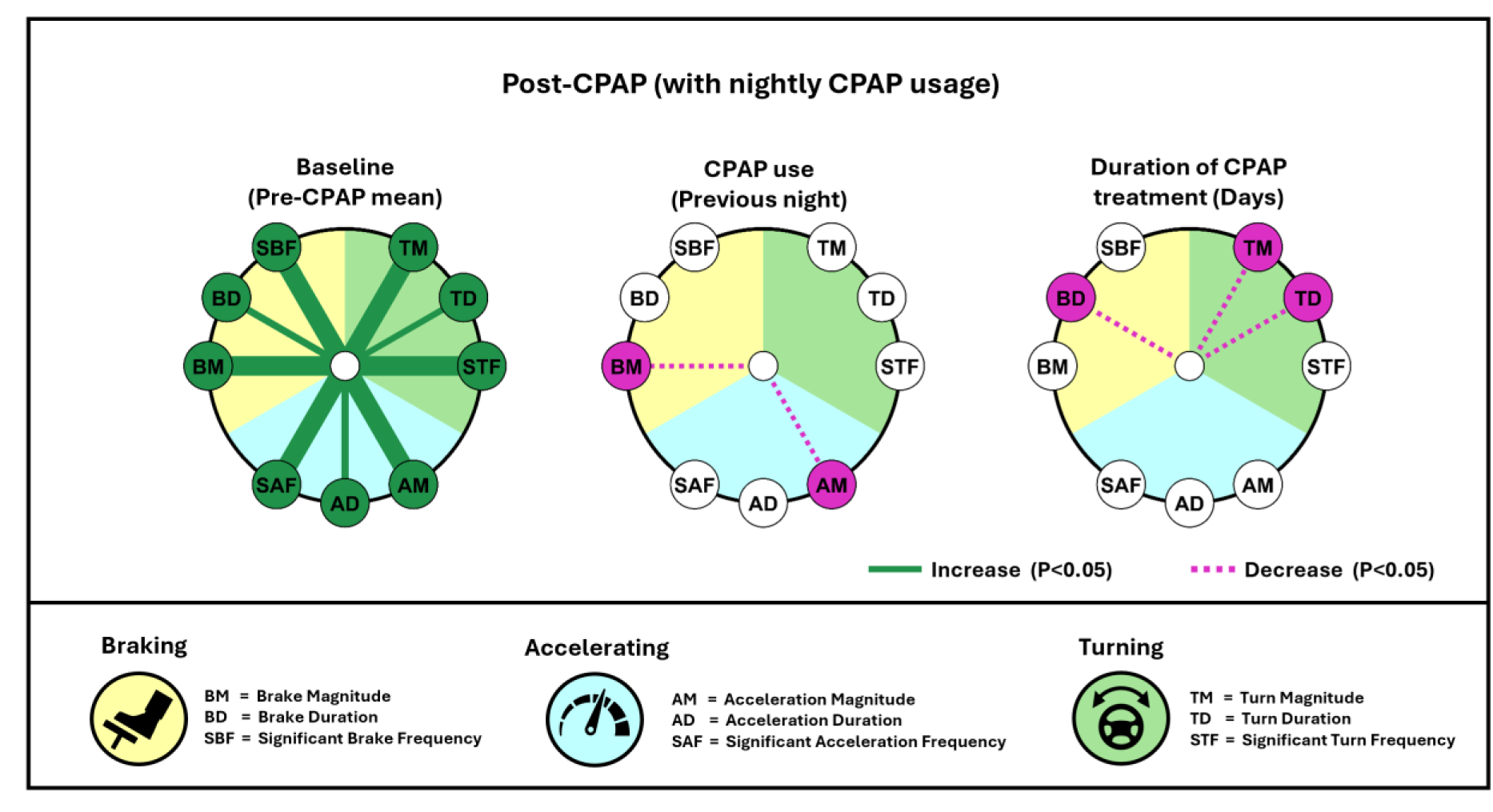
Radial significance plots showing that baseline (Pre-CPAP) driving strongly predicts Post-CPAP driving outcomes (9/9), with additional associations for previous night’s CPAP use (2/9) and duration of CPAP treatment (3/9). Radial significance plots display linear mixed-effect (LME) model results testing the effect of Pre-CPAP driving, CPAP use, duration of CPAP treatment on driving performance. Each circle reflects independent fixed effects within models. Outer markers reflect driving measures tested grouped by type: (Braking (yellow): BM = brake magnitude, BD = brake duration, SBF = significant brake frequency, Acceleration (blue): AM = acceleration magnitude, AD = acceleration duration, SAF = significant acceleration frequency and Turning (green): TM = turn magnitude, TD = turn duration, STF = significant turn frequency). Presence of a line indicates significance, green (increase), purple (decrease), width indicates effect size, cut at a maximum of 1.2 and a minimum of 0.05.

Time into trip and trip duration were significant predictors for most outcomes, reinforcing the importance of accounting for trip structure in modelling real-world driving behaviour.

To assess whether short-term adherence history strengthened these associations, models were re-estimated using mean CPAP use across the preceding 2-3 nights. Compared with single-night use, multi-night averages showed additional associations with driving behaviour. With the two-night average, CPAP use was additionally associated with reduced turn duration (P < 0.001, ES = −0.05), while the three-night average was additionally associated with both reduced turn duration (P < 0.001, ES = −0.05) and lower significant acceleration frequency (P < 0.05, ES = −0.17) (supplementary tables e and f). Baseline driving and treatment-duration effects were unchanged. Although effect sizes were small, these findings suggest that recent multi-night adherence relates more consistently to real-world driving behaviour than single-night CPAP use.

### Post-CPAP Interaction Effects: CPAP usage affects driving differently depending on Baseline (Pre-CPAP) driving style

Given the strong main effects of baseline (Pre-CPAP) driving behaviour, interaction analyses were conducted to test whether baseline driving style moderated the effects of CPAP use and treatment duration on Post-CPAP driving. Significant baseline × prior-night CPAP use interactions were observed for four outcomes: brake duration (P < 0.001, ES = −0.09), significant brake frequency (P < 0.001, ES = −0.30), acceleration magnitude (P < 0.001, ES = −0.07), and significant acceleration frequency (P < 0.001, ES = −0.34). Considering the strong positive main effects of baseline driving, these negative interactions indicate convergence of driving behaviour with increasing CPAP use, such that differences between low- and high-baseline drivers attenuated with greater adherence.

For context, driving behaviour in clinic attendees without OSA (ODI <5) or excessive daytime sleepiness (ESS <11) was summarised using participant-level means. Control values fell within the trend bands defined by baseline groups across the CPAP adherence range. For braking outcomes, both baseline groups converged toward control-like performance with increasing CPAP use. For acceleration outcomes, low-baseline drivers were already comparable to controls, while high-baseline drivers progressively shifted toward control levels with greater adherence (figure 4; supplementary table g). See supplementary text d for additional interaction results.

**Figure 4:**
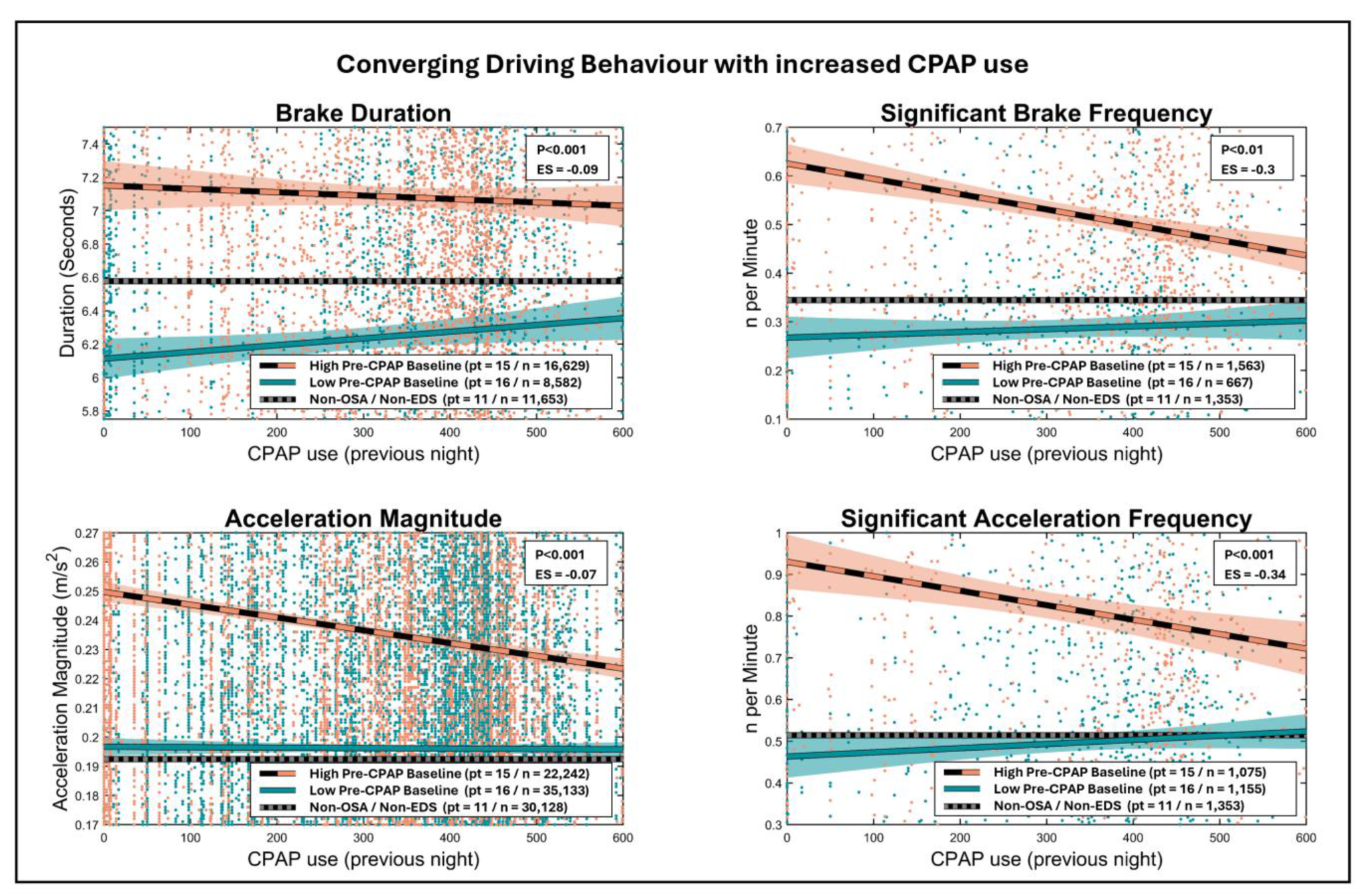
Scatter plots illustrating the dose-response relationship between CPAP use and (4/9) driving outcomes, showing modulation by baseline (Pre-CPAP) driving style. Ordinary least squares (OLS) trend lines are displayed for the four driving measures with significant CPAP usage × baseline interaction effects. Orange markers and long-dash lines, reflects data from participants with the highest baseline scores, turquoise markers and lines represent participants with low baseline scores. Shaded areas denote 95% confidence interval of the OLS trend lines. Short-dash grey lines represent the mean of control-like participants (Non-OSA / Non-EDS) averages. P value and effect sizes (ES), correspond to results from the linear mixed-effect (LME) models.

## Discussion

In a representative cohort of patients, we examined how CPAP treatment alters real-world driving behaviour in patients with OSA. Across more than seven thousand trips, CPAP use produced measurable changes in six of nine driving metrics, reflecting a sharper and less aggressive style. Pre-CPAP (baseline) behaviour was strongly related to Post-CPAP behaviour, and nightly adherence predicted convergence towards control like behaviour that depended on each person’s baseline performance. Drivers who were initially either overly aggressive or overly cautious both moved toward control-like behaviour as adherence increased. These findings show that OSA related driving impairment can appear in different behavioural forms and that CPAP normalises these patterns with increased use.

### Convergence in driving styles as modulated by CPAP usage

Patients with distinct Pre-CPAP braking and acceleration styles showed different dose–response relationships between adherence and driving outcomes, indicating that CPAP influences behavioural regulation as well as average performance. This aligns with the recognised heterogeneity of OSA symptoms (Ye et al., 2014; Gomez-Pilar et al., 2023) and suggests that impairment reflects dysregulated control rather than uniform slowing or speeding. Unlike common simulator outcomes such as reaction time or lane deviation, real-world driving requires balance, such that both excessive and insufficient manoeuvre intensity or duration may be suboptimal and potentially increase crash risk.

To our knowledge, these findings are the first to demonstrate convergence in real-world driving style as a function of CPAP use. Previous large cohort studies have shown that greater mean CPAP adherence is associated with fewer near-misses and collisions (Karimi et al., 2015; Coelho et al., 2024), but do not address how driving behaviour itself changes. Simulator studies have reported substantial inter-individual variability in impairment (George et al., 1996; Vakulin et al., 2011) and in vulnerability to performance decline with extended wakefulness (Vakulin et al., 2022). Consistent with this, the US Department of Transportation describes both aggressive (e.g. speeding) and excessively cautious (e.g. wide turns, markedly reduced speed) behaviours as indicators of impaired driving (Department of Transportation, 2010), highlighting that impairment does not manifest uniformly.

Our results show that CPAP treatment modulates these distinct driving styles, promoting convergence toward more stable and adaptive patterns of behaviour. Whether overly aggressive and overly cautious styles confer equivalent crash risk remains unclear; excessively cautious driving may reflect a conscious compensatory strategy among drivers aware of their fatigue. Importantly, the identification of dose–response relationships between nightly CPAP use and driving outcomes supports the sensitivity of telematics-based metrics for capturing real-world functional change. The stronger interaction effects observed for event-frequency measures further suggest that frequency-based metrics may be particularly informative for assessing on-treatment driving impairment in OSA

### Pre vs Post-CPAP driving

Our Pre- versus Post-CPAP analysis identified significant changes in six of nine driving outcomes, consistent with prior driving simulator studies reporting improved performance following CPAP initiation (Hack et al., 2001; Turkington et al., 2004; Orth et al., 2005; Vakulin et al., 2011). Reductions in acceleration frequency, turn magnitude, and turn frequency indicate a shift toward less aggressive driving, while shorter braking, acceleration, and turning durations suggest sharper, more decisive manoeuvres at the group level. Except for turn frequency, effect sizes were small and varied across individuals, limiting immediate clinical interpretability. However, given the complexity of real-world driving and the multiple factors that influence crash risk, even modest behavioural changes may translate into meaningful population-level risk reductions. Importantly, our findings also show that CPAP-related changes depend on Pre-treatment driving style, indicating that simple group contrasts may obscure clinically relevant heterogeneity in treatment response.

### The importance of Baseline (Pre-CPAP) driving

Baseline driving behaviour predicted all nine outcomes and showed particularly strong associations with magnitude and frequency measures, indicating that Pre-treatment performance was the strongest determinant of Post-CPAP driving behaviour among the covariates examined. This aligns with recent cohort data showing that patients with a prior history of driving incidents remain at substantially higher risk of near-misses and collisions after CPAP initiation compared with those without such history (Coelho et al., 2024). Together, these findings suggest that patients with recent near-miss or collision events before starting CPAP may remain at elevated risk in the short to mid-term and therefore warrant closer clinical monitoring and explicit counselling regarding driving safety and the importance of treatment adherence. From a public health perspective, this also raises the question of whether driving guidance and monitoring strategies should more explicitly account for prior incident history when assessing risk after treatment initiation.

### Compliance windows

Given the limited evidence on how many consecutive nights of CPAP use are required to influence driving performance, we compared adherence windows of one, two, and three nights. Associations with driving outcomes were stronger for two- and three-night averages than for prior-night use alone, suggesting that a short multi-night window may better capture adherence-related behavioural change. This is consistent with simulator studies showing improvements over the first two nights of CPAP use before stabilisation (Turkington et al., 2004; Orth et al., 2005). However, the incremental effects observed with longer windows were small and unlikely to add substantial clinical value. Evidence from sleep restriction studies similarly indicates that a single night of recovery sleep may be insufficient for full restoration of driving-related performance (Dinges et al., 1997; Banks et al., 2010; Yamazaki et al., 2021). In contrast, longer-term adherence averages used in prior studies (e.g. ≥90 days (Coelho et al., 2024) or approximately three months (Vakulin et al., 2011)) may obscure the effects of short-term adherence fluctuations. Together, these findings support the use of nightly or short-window CPAP adherence data when examining dose–response relationships with driving behaviour and highlight the need for future work on the impact of irregular adherence patterns.

### From group effects to personalised monitoring

The present findings confirm that CPAP influences real-world driving performance and extend prior work by characterising behavioural normalisation with treatment. While large cohort studies show that adherent CPAP users have fewer collisions and near misses (George, 2001; Barbé et al., 2007; Karimi et al., 2015; Coelho et al., 2024), they cannot reveal intermediate behavioural mechanisms. Here, increasing adherence was associated with reduced extremes of manoeuvre intensity, providing a plausible behavioural pathway linking treatment use to safer driving outcomes.

Smartphone telematics offers a scalable method to bridge clinical monitoring and everyday behaviour. Passive data collection could support tracking of recovery trajectories and residual risk, complementing CPAP device adherence data. Behaviour-based assessment may also support clinical and regulatory decisions, which currently rely largely on diagnosis and self-report (Hartenbaum et al., 2006; Evans et al., 2017).

### Strengths, limitations and future directions

Key strengths of this study include its ecologically valid assessment of real-world driving behaviour, the large volume of data collected (>450,000 driving events), and integration within the routine clinical pathway from diagnosis to treatment. Mixed-effects models accounted for participant-level variability and trip structure. Limitations include unequal numbers of trips per participant and a small control group, which may limit generalisability.

Future work should test whether telematics-derived driving metrics predict verified crash outcomes and replicate these findings in larger and more diverse samples. Studies are also needed to determine phenotype-specific adherence thresholds, the number of consecutive nights of adequate CPAP use required for behavioural recovery, and the impact of inconsistent adherence. Such evidence would directly inform clinical advice and driving safety guidance.

## Conclusion

We have assessed real-world driving performance Pre and Post-CPAP initiation for OSA, as recommended as a priority by the European Respiratory Society (Bonsignore et al., 2021). Our results showed that driving behaviour in OSA is heterogeneously impaired, and CPAP use promotes convergence towards control-like performance. As adherence increases, both overly aggressive and overly cautious driving patterns move towards typical behaviour. These findings refine understanding of how OSA influences everyday driving and how treatment restores behavioural regulation. Smartphone telematics provide an affordable and scalable method to monitor these changes and may become an important tool for personalised assessment of risk, treatment efficacy monitoring, and driving safety in sleep and respiratory clinics.

## Data Availability

The data that support the findings of this study are not publicly available due to restrictions imposed by ethical approvals and participant consent. Reasonable requests for access to anonymised data may be considered by the corresponding author, subject to institutional approval.

## Acknowledgments

We would like to express appreciation to all the participants who volunteered to take part in the study, Royal Papworth Charity for financial support, and all departments across the University of Cambridge and Royal Papworth Hospital for facilitating study set-up and delivery.

## Supplementary Material

### Supplementary Figures

**Supplementary Figure a:**
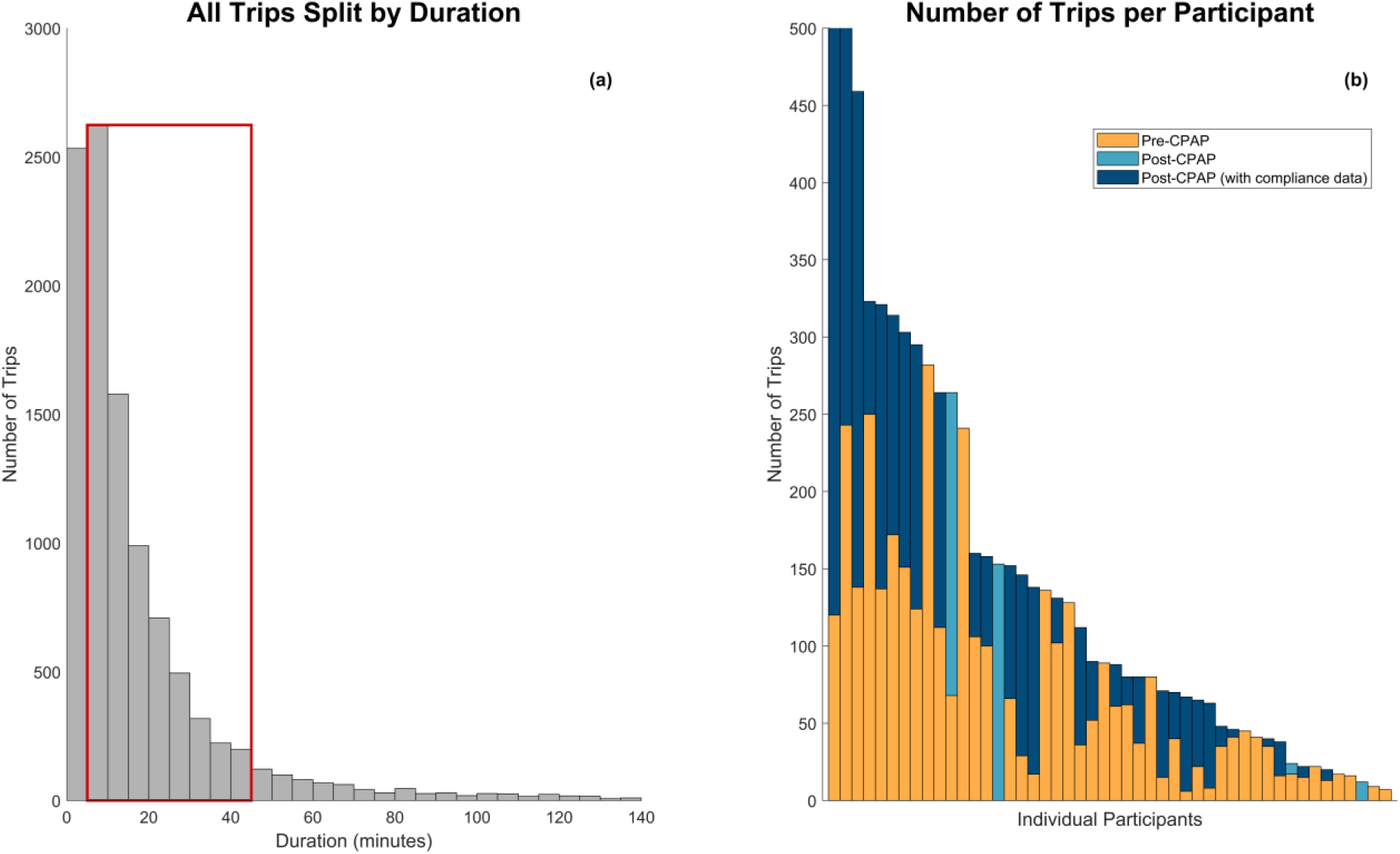
a) Histogram showing the number of trips collected split by 5 minute intervals, cut at 140 minutes on the x axis. The red box contains trips with a duration between 5-45 minutes, used for analysis. B) Bar chart showing the number of trips completed per participant, cut at 500 on the y axis. Yellow (bottom where split) reflects trips completed Pre-CPAP, light blue Post-CPAP, and dark blue Post-CPAP with compliance data.

**Supplementary Figure b:**
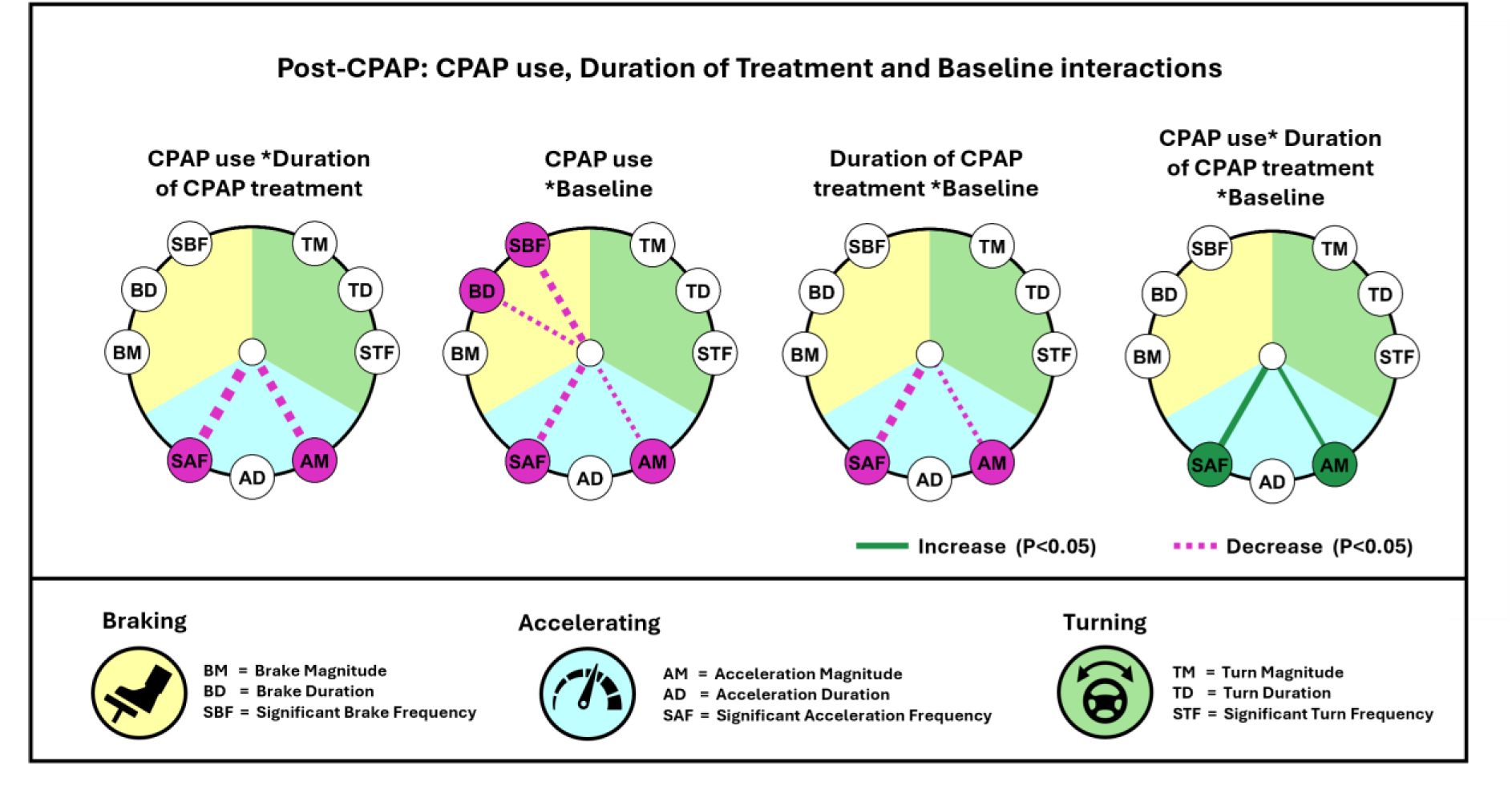
Radial significance plots showing interaction results of CPAP use, duration of CPAP treatment and baseline (Pre-CPAP) driving. Radial significance plots display linear mixed-effect (LME) model results testing the effect of Pre-CPAP driving, CPAP use, duration of CPAP treatment on driving performance. Each circle reflects independent fixed effects within models. Outer markers reflect driving measures tested grouped by type: (Braking (yellow): BM = brake magnitude, BD = brake duration, SBF = significant brake frequency, Acceleration (blue): AM = acceleration magnitude, AD = acceleration duration, SAF = significant acceleration frequency and Turning (green): TM = turn magnitude, TD = turn duration, STF = significant turn frequency). Presence of a line indicates significance, green (increase), purple (decrease), width indicates effect size, cut at a maximum of 1.2 and a minimum of 0.05.

### Supplementary Text

#### Supplementary Text a: Supplementary Methods

All patients referred to the sleep service during the recruitment window received an invitation letter alongside routine appointment correspondence. The invitation included a QR code linking to the study information sheet and an online screening questionnaire. Following expression of interest, a member of the research team contacted potential participants by telephone to explain study procedures, confirm eligibility, and arrange enrolment. Informed consent was obtained using the DocuSign online platform. Participants were guided through installation and activation of the smartphone application by the study team. Patients with negative diagnostic testing for OSA or who did not commence CPAP therapy were excluded from the primary analysis. CPAP adherence data were obtained directly from device memory cards following the Post-treatment monitoring period.

At the study site, the majority of patients undergo diagnostic assessment using home pulse oximetry. In this cohort, 41 participants (85.4%) did not require further testing beyond oximetry. For analytic simplicity, ODI and AHI values were combined under a single severity label (ODI), reflecting the primary screening metric used in routine care.

#### Supplementary Text b: Data retention and preprocessing

Nineteen thousand three hundred and thirty journeys were recorded from 98 participants. Two participants were excluded due to absence of recorded driving data. Short-duration trips (<5 minutes; 24.2% of journeys) were excluded as they were overrepresented and less informative of sustained driving behaviour. Long journeys (>45 minutes; 7.7%) were infrequent and contributed by a small number of participants and were therefore excluded to reduce disproportionate influence. Trips occurring more than 100 days after CPAP initiation were also excluded. After exclusions, the final dataset comprised 7,134 trips from 48 participants (mean 149 trips per participant; range 7–810). Participant-level trip distributions are shown in supplementary figure a.

Nightly CPAP adherence data were extracted from device memory cards and matched to next-day driving behaviour, yielding a compliance-linked dataset of 2,232 trips. A control dataset was compiled from patients who underwent oximetry screening, and were OSA negative (ODI < 5) and did not have EDS (ESS < 11), producing a dataset of 1,353 trips from 11 participants (mean ODI: 2.5 [range 0.1-4.3]; mean ESS: 7.1 [0-10]; 55% female; mean age: 56 [39-69]).

#### Supplementary Text c: Supplementary Statistical Models

Statistical values were calculated using linear mixed-effect (LME) models. Time into trip (not for frequency measures) and trip duration were always included as fixed effects, and individual participants were included as random effects, controlling for different number of journeys undertaken and other potential confounding factors across participants. Each analysis contained six or nine models (one for each driving measure, except when time into trip was measured and frequency measures could not be tested). Effect size (ES) was calculated by the LME fixed effect estimate / residual standard deviation. For independent effects, continuous variables ES’s were scaled by two standard deviations, to aid interpretability across the distribution, and comparison with categorical ES’s, in line with recommendations (Gelman, 2008). However, baseline measures reflecting individual participants Pre-CPAP driving were left unscaled when tested independently. This was due to baseline already being averaged, reducing within-participant variability, and applying additional scaling could distort the practical meaning of the resulting ES’s. In contrast, when baseline was part of an interaction term (e.g. CPAP use and baseline interactions), we did apply two standard deviation scaling. This ensured that resulting ES’s from interaction results reflect meaningful combined changes across the observed range of each variable, and compatibility with other continuous predictors. P values were adjusted using a Bonferroni correction, with the correction factor fixed at nine (number of driving measures) of all models, including frequency measures. All analysis was conducted in MATLAB (version R2024a).

#### Supplementary Text d: Further Interaction Results

Additional interaction analyses showed that CPAP use × duration of treatment was significant for 2/9 outcomes: decreased acceleration magnitude (P < 0.01, ES: -0.43), and significant acceleration frequency (P < 0.05, ES: -0.61). Indicating that the association of prior-night CPAP use with acceleration behaviour diminishes as time on treatment increases. Treatment duration × baseline interaction was also significant for the same 2/9 outcomes: decreased acceleration magnitude (P < 0.05, ES: -0.1), and significant acceleration frequency (P < 0.05, ES: -0.46). Suggesting that the effect of treatment duration on acceleration behaviours also depends on Pre-CPAP driving. The three-way interaction of CPAP use × treatment duration × baseline were significant for 2/9 outcomes: increased acceleration magnitude (P < 0.001, ES: 0.05), and significant acceleration frequency (P < 0.01, ES: 0.24) (supplementary figure b; supplementary table g).

### Supplementary Tables

**Supplementary Table a** : Motion signals from smartphone accelerometers and gyroscopes were processed to detect discrete braking, acceleration, and turning events. For each event, both magnitude and duration were quantified. Events exceeding predefined magnitude thresholds were classified as “significant,” and the frequency of significant events per minute of driving was calculated. In total, nine telematics-derived driving metrics were generated and used in statistical analyses.

**Supplementary Table a:** Driving Performance Measures.

| Supplementary Table a: Driving Performance Measures |  |  |
| --- | --- | --- |
| Measure | Abbreviation | Description |
| Brake Magnitude | BM | The force of the brake event in m/s<sup>2</sup> |
| Brake Duration | BD | The length of the brake event in seconds |
| Significant Brake Frequency | **SBF** | Number of times a brake event exceeded a magnitude of 0.31 m/s<sup>2</sup> per minute of driving for a single trip |
| Acceleration Magnitude | **AM** | The force of the magnitude event in m/s<sup>2</sup> |
| Acceleration Duration | **AD** | The length of the magnitude event in seconds |
| Significant Acceleration Frequency | **SAF** | Number of times an acceleration event exceeded a magnitude of 0.23 m/s<sup>2</sup> per minute of driving for a single trip |
| Turn Magnitude | **TM** | The force of the turn event in m/s<sup>2</sup> |
| Turn Duration | **TD** | The length of a turn event in seconds |
| Significant Turn Frequency | **STF** | Number of times a turn event exceeded a magnitude of 0.36 m/s<sup>2</sup> per minute of driving for a single trip |

**Supplementary Table b:** Data used for each analysis.

| Data | Analysis |  |
| --- | --- | --- |
|  | Pre and Post-CPAP | Post-CPAP (with nightly usage) |
| n participants | 48 | 31 |
| n trips | 7,134 | 2,232 |
| n brake events | 83,734 | 25,211 |
| n acceleration events | 188,881 | 57,375 |
| n turn events | 186,016 | 55,484 |
| n total events | 458,631 | 138,070 |
| Total driving time (hrs) | 1,872 | 572 |
| Mean trip duration (mins) | 15.7 | 15.4 |
| Diagnostic data |  |  |
| Mean ODI (SD / range) | 25.4 (17.3 / 5.3-84.2) | 26.2 (18.1 / 5.9-84.2) |
| Mean ESS (SD / range) | 13.6 (5.2 / 1-24) | 14.4 (5.2 / 5-24) |
| Demographics |  |  |
| Mean age years (SD / range) | 53.5 (13.4 / 30-77) | 57 (12 / 36-77) |
| Sex F/M (% female) | 13/35 (37.1%) | 8/23 (25.1%) |
| Mean BMI kg/m<sup>2</sup> (SD / range) | 34.8 (8.6 / 23-58) | 34.7 (9.1 / 23-58) |

**Supplementary Table c:**
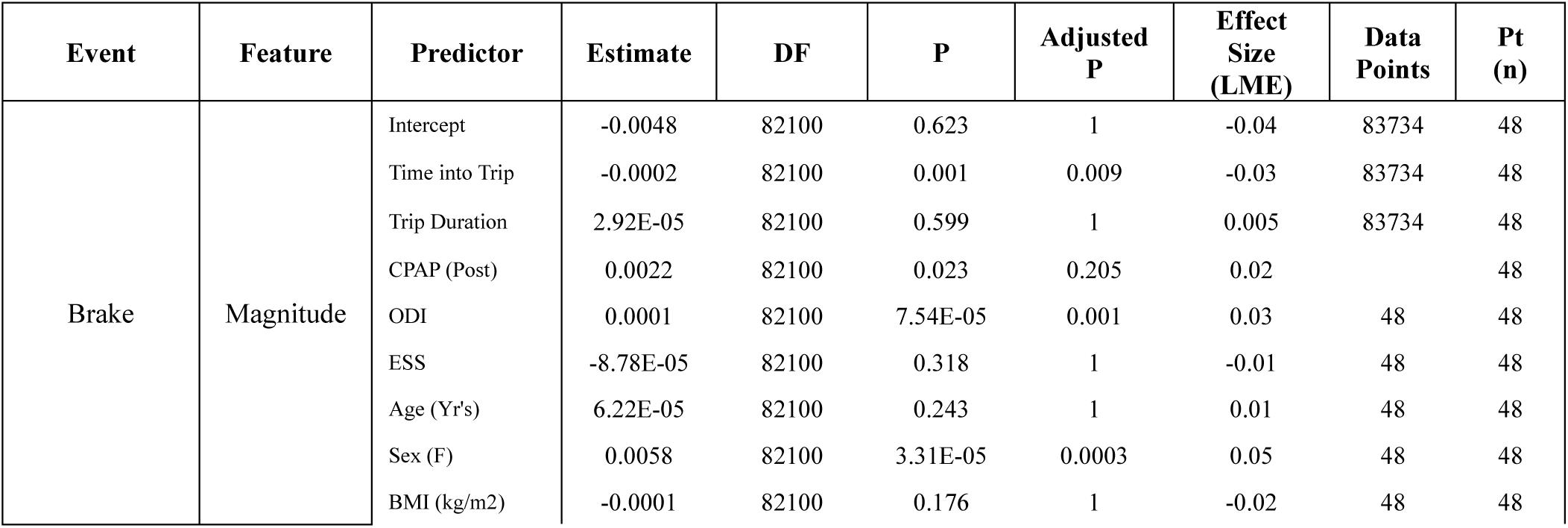
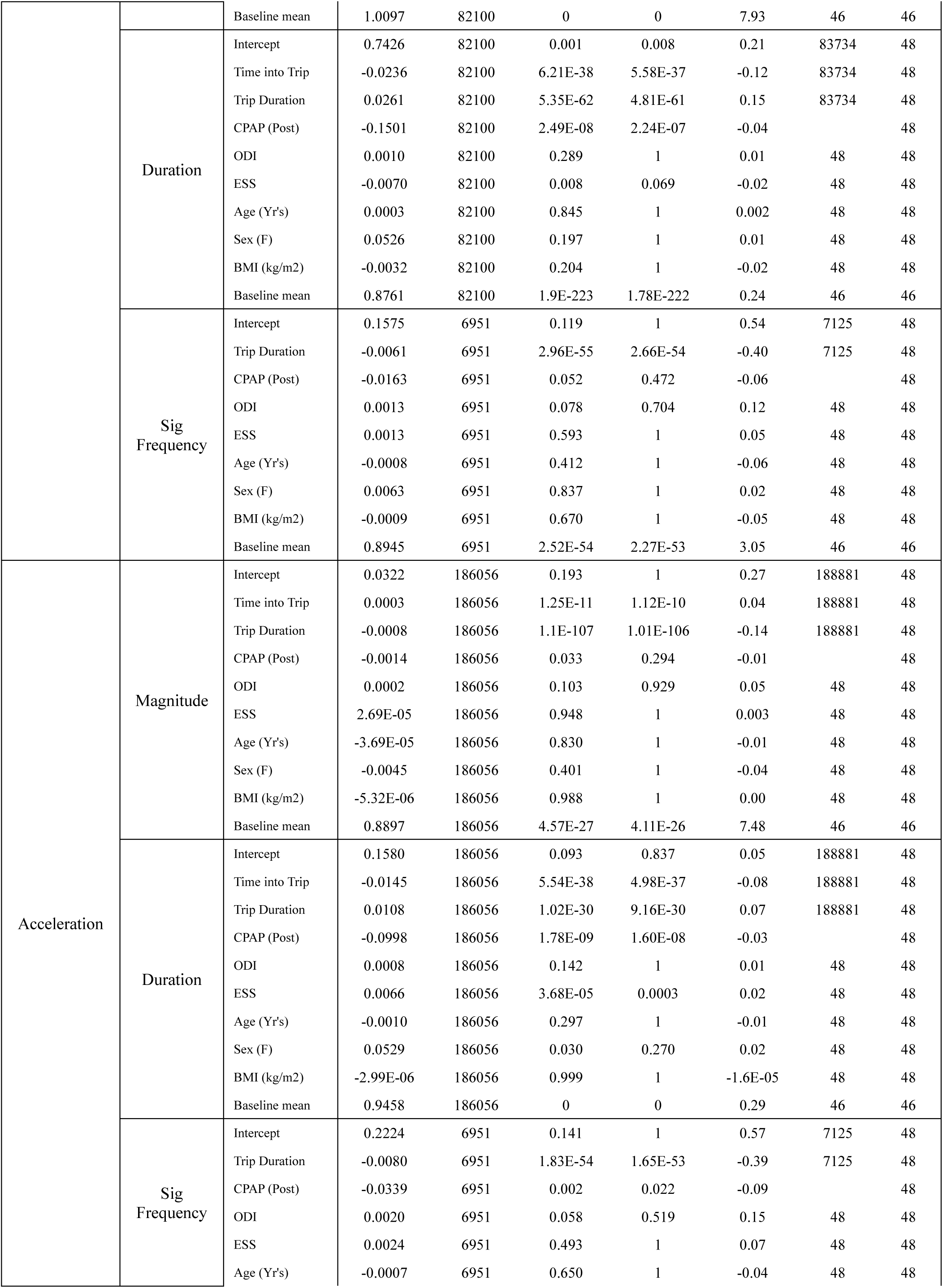
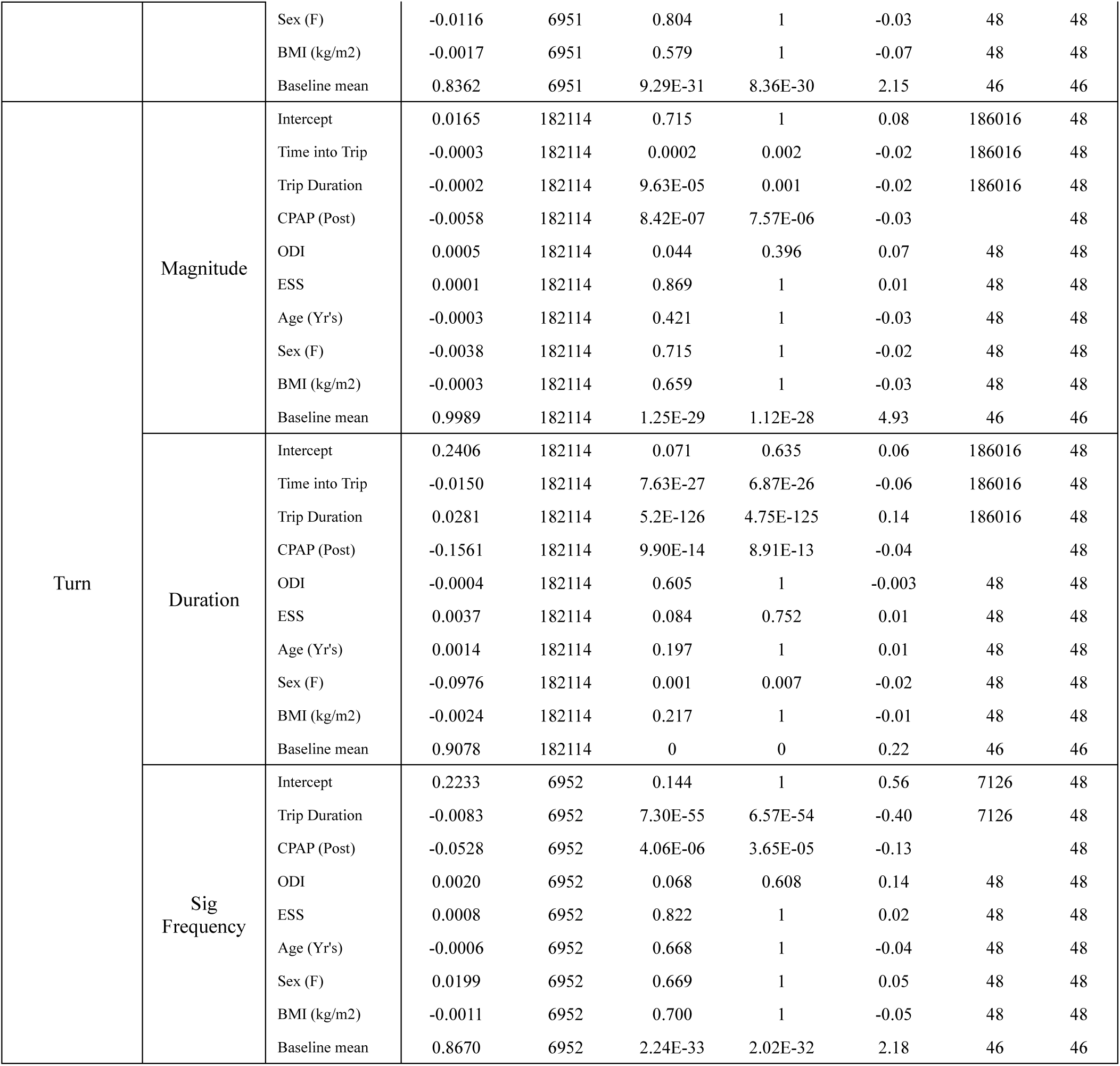
Pre and Post CPAP LME Results.

**Supplementary Table d:**
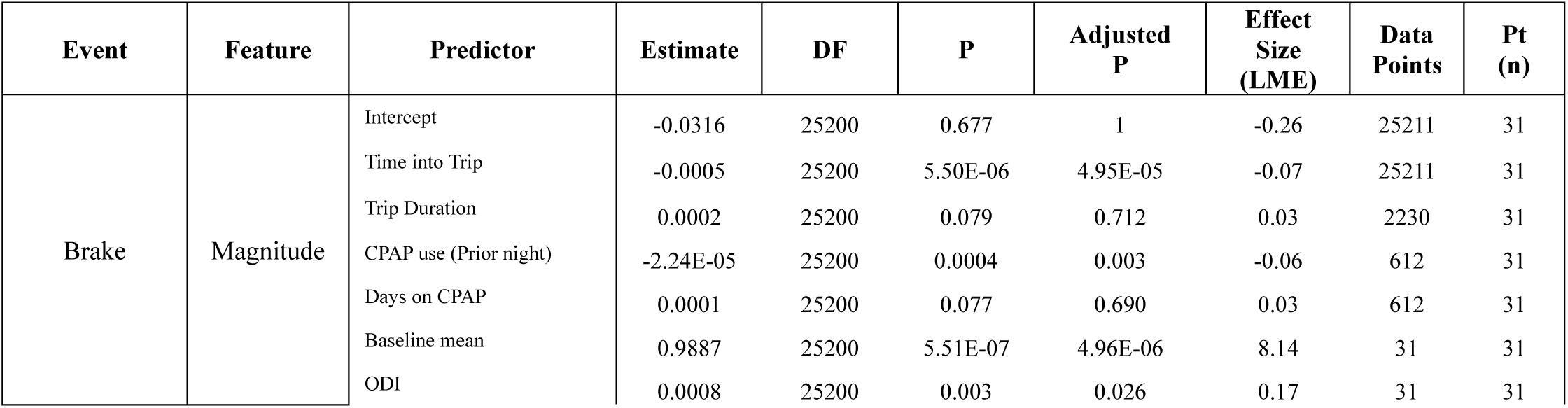
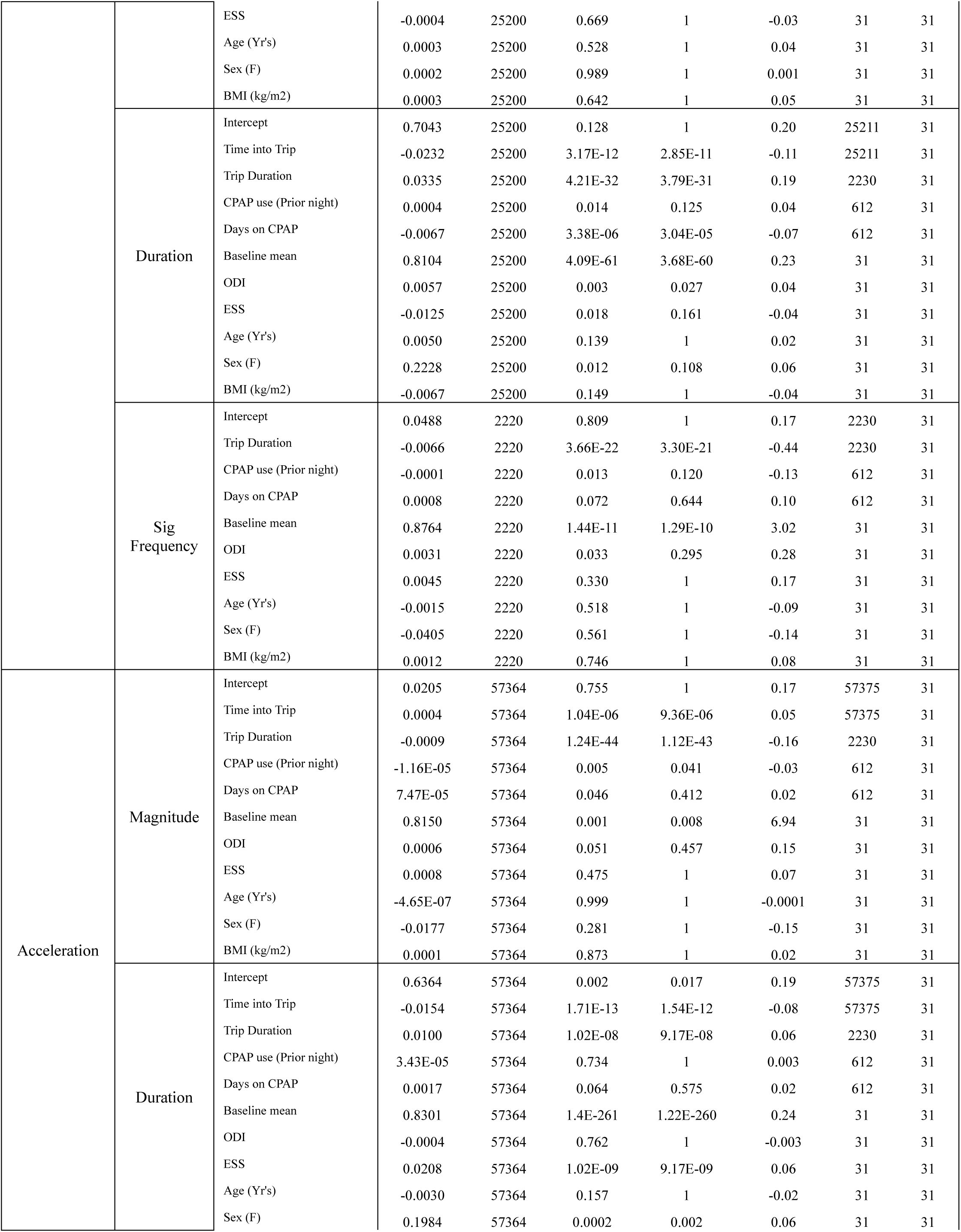
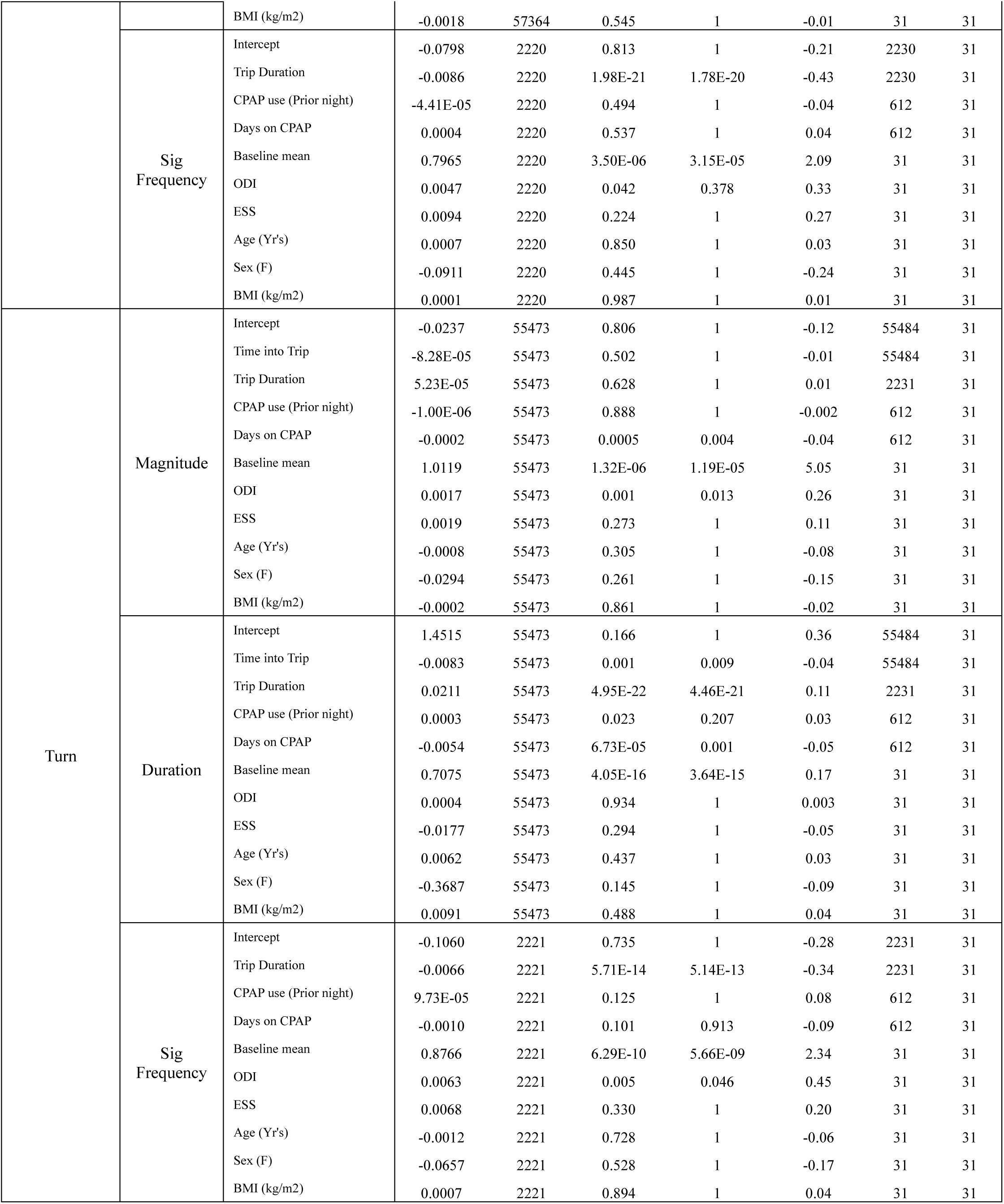
CPAP Compliance Results (Prior night’s usage)

**Supplementary Table e:** CPAP Compliance Results (Previous 2 night’s mean usage)

| Event | Feature | Predictor | Estimate | DF | P | Adjusted P | Effect Size (LME) | Data Points | Pt (n) |
| --- | --- | --- | --- | --- | --- | --- | --- | --- | --- |
| Brake | Magnitude | Intercept | -0.0277 | 25200 | 0.721 | 1 | -0.23 | 25211 | 31 |
|  |  | Time into Trip | -0.0005 | 25200 | 5.56E-06 | 5.00E-05 | -0.07 | 25211 | 31 |
|  |  | Trip Duration | 0.0002 | 25200 | 0.094 | 0.843 | 0.03 | 2230 | 31 |
|  |  | CPAP use (2 night mean) | -2.83E-05 | 25200 | 8.87E-05 | 0.001 | -0.07 | 612 | 31 |
|  |  | Days on CPAP | 0.0001 | 25200 | 0.044 | 0.392 | 0.04 | 612 | 31 |
|  |  | Baseline mean | 0.9833 | 25200 | 1.04E-06 | 9.35E-06 | 8.10 | 31 | 31 |
|  |  | ODI | 0.0008 | 25200 | 0.003 | 0.029 | 0.18 | 31 | 31 |
|  |  | ESS | -0.0004 | 25200 | 0.647 | 1 | -0.04 | 31 | 31 |
|  |  | Age (Yr's) | 0.0002 | 25200 | 0.572 | 1 | 0.03 | 31 | 31 |
|  |  | Sex (F) | 0.0004 | 25200 | 0.976 | 1 | 0.003 | 31 | 31 |
|  |  | BMI (kg/m2) | 0.0003 | 25200 | 0.625 | 1 | 0.05 | 31 | 31 |
|  | Duration | Intercept | 0.7276 | 25200 | 0.116 | 1 | 0.20 | 25211 | 31 |
|  |  | Time into Trip | -0.0233 | 25200 | 3.03E-12 | 2.73E-11 | -0.11 | 25211 | 31 |
|  |  | Trip Duration | 0.0336 | 25200 | 3.34E-32 | 3.01E-31 | 0.19 | 2230 | 31 |
|  |  | CPAP use (2 night mean) | 0.0003 | 25200 | 0.078 | 0.698 | 0.03 | 612 | 31 |
|  |  | Days on CPAP | -0.0065 | 25200 | 5.87E-06 | 5.28E-05 | -0.07 | 612 | 31 |
|  |  | Baseline mean | 0.8105 | 25200 | 3.93E-61 | 3.54E-60 | 0.23 | 31 | 31 |
|  |  | ODI | 0.0058 | 25200 | 0.003 | 0.023 | 0.04 | 31 | 31 |
|  |  | ESS | -0.0133 | 25200 | 0.012 | 0.105 | -0.04 | 31 | 31 |
|  |  | Age (Yr's) | 0.0047 | 25200 | 0.159 | 1 | 0.02 | 31 | 31 |
|  |  | Sex (F) | 0.2198 | 25200 | 0.013 | 0.119 | 0.06 | 31 | 31 |
|  |  | BMI (kg/m2) | -0.0060 | 25200 | 0.194 | 1 | -0.03 | 31 | 31 |
|  | Sig Frequency | Intercept | 0.0637 | 2220 | 0.756 | 1 | 0.22 | 2230 | 31 |
|  |  | Trip Duration | -0.0067 | 2220 | 2.66E-22 | 2.39E-21 | -0.44 | 2230 | 31 |
|  |  | CPAP use (2 night mean) | -0.0002 | 2220 | 0.006 | 0.050 | -0.16 | 612 | 31 |
|  |  | Days on CPAP | 0.0009 | 2220 | 0.047 | 0.422 | 0.11 | 612 | 31 |
|  |  | Baseline mean | 0.8781 | 2220 | 2.33E-11 | 2.10E-10 | 3.03 | 31 | 31 |
|  |  | ODI | 0.0031 | 2220 | 0.032 | 0.289 | 0.29 | 31 | 31 |
|  |  | ESS | 0.0042 | 2220 | 0.365 | 1 | 0.16 | 31 | 31 |
|  |  | Age (Yr's) | -0.0017 | 2220 | 0.481 | 1 | -0.10 | 31 | 31 |
|  |  | Sex (F) | -0.0387 | 2220 | 0.583 | 1 | -0.13 | 31 | 31 |
|  |  | BMI (kg/m2) | 0.0013 | 2220 | 0.721 | 1 | 0.09 | 31 | 31 |
| Acceleration | Magnitude | Intercept | 0.0237 | 57364 | 0.720 | 1 | 0.20 | 57375 | 31 |
|  |  | Time into Trip | 0.0004 | 57364 | 9.94E-07 | 8.95E-06 | 0.05 | 57375 | 31 |
|  |  | Trip Duration | -0.0009 | 57364 | 4.98E-45 | 4.48E-44 | -0.16 | 2230 | 31 |
|  |  | CPAP use (2 night mean) | -1.82E-05 | 57364 | 0.0001 | 0.001 | -0.04 | 612 | 31 |
|  |  | Days on CPAP | 8.86E-05 | 57364 | 0.019 | 0.171 | 0.03 | 612 | 31 |
|  |  | Baseline mean | 0.8117 | 57364 | 0.001 | 0.009 | 6.92 | 31 | 31 |
|  |  | ODI | 0.0006 | 57364 | 0.051 | 0.456 | 0.15 | 31 | 31 |
|  |  | ESS | 0.0007 | 57364 | 0.499 | 1 | 0.06 | 31 | 31 |
|  |  | Age (Yr's) | -2.40E-05 | 57364 | 0.961 | 1 | -0.004 | 31 | 31 |
|  |  | Sex (F) | -0.0175 | 57364 | 0.291 | 1 | -0.15 | 31 | 31 |
|  |  | BMI (kg/m2) | 0.0002 | 57364 | 0.850 | 1 | 0.03 | 31 | 31 |
|  | Duration | Intercept | 0.6459 | 57364 | 0.002 | 0.015 | 0.19 | 57375 | 31 |
|  |  | Time into Trip | -0.0154 | 57364 | 1.72E-13 | 1.55E-12 | -0.08 | 57375 | 31 |
|  |  | Trip Duration | 0.0100 | 57364 | 1.18E-08 | 1.07E-07 | 0.06 | 2230 | 31 |
|  |  | CPAP use (2 night mean) | -1.57E-05 | 57364 | 0.889 | 1 | -0.001 | 612 | 31 |
|  |  | Days on CPAP | 0.0017 | 57364 | 0.053 | 0.478 | 0.02 | 612 | 31 |
|  |  | Baseline mean | 0.8310 | 57364 | 2.1E-261 | 1.87E-260 | 0.24 | 31 | 31 |
|  |  | ODI | -0.0003 | 57364 | 0.818 | 1 | -0.002 | 31 | 31 |
|  |  | ESS | 0.0204 | 57364 | 2.11E-09 | 1.90E-08 | 0.06 | 31 | 31 |
|  |  | Age (Yr's) | -0.0031 | 57364 | 0.143 | 1 | -0.02 | 31 | 31 |
|  |  | Sex (F) | 0.1969 | 57364 | 0.0003 | 0.002 | 0.06 | 31 | 31 |
|  |  | BMI (kg/m2) | -0.0015 | 57364 | 0.608 | 1 | -0.01 | 31 | 31 |
|  | Sig Frequency | Intercept | -0.0512 | 2220 | 0.880 | 1 | -0.13 | 2230 | 31 |
|  |  | Trip Duration | -0.0086 | 2220 | 1.56E-21 | 1.40E-20 | -0.43 | 2230 | 31 |
|  |  | CPAP use (2 night mean) | -0.0001 | 2220 | 0.117 | 1 | -0.09 | 612 | 31 |
|  |  | Days on CPAP | 0.0005 | 2220 | 0.430 | 1 | 0.05 | 612 | 31 |
|  |  | Baseline mean | 0.7899 | 2220 | 4.92E-06 | 4.43E-05 | 2.08 | 31 | 31 |
|  |  | ODI | 0.0048 | 2220 | 0.041 | 0.372 | 0.34 | 31 | 31 |
|  |  | ESS | 0.0090 | 2220 | 0.246 | 1 | 0.26 | 31 | 31 |
|  |  | Age (Yr's) | 0.0005 | 2220 | 0.897 | 1 | 0.02 | 31 | 31 |
|  |  | Sex (F) | -0.0898 | 2220 | 0.455 | 1 | -0.24 | 31 | 31 |
|  |  | BMI (kg/m2) | 0.0004 | 2220 | 0.942 | 1 | 0.02 | 31 | 31 |
| Turn | Magnitude | Intercept | -0.0229 | 55473 | 0.813 | 1 | -0.11 | 55484 | 31 |
|  |  | Time into Trip | -8.28E-05 | 55473 | 0.502 | 1 | -0.01 | 55484 | 31 |
|  |  | Trip Duration | 5.20E-05 | 55473 | 0.630 | 1 | 0.01 | 2231 | 31 |
|  |  | CPAP use (2 night mean) | -2.64E-06 | 55473 | 0.753 | 1 | -0.004 | 612 | 31 |
|  |  | Days on CPAP | -0.0002 | 55473 | 0.001 | 0.005 | -0.04 | 612 | 31 |
|  |  | Baseline mean | 1.0111 | 55473 | 1.38E-06 | 1.24E-05 | 5.04 | 31 | 31 |
|  |  | ODI | 0.0017 | 55473 | 0.001 | 0.013 | 0.26 | 31 | 31 |
|  |  | ESS | 0.0019 | 55473 | 0.275 | 1 | 0.11 | 31 | 31 |
|  |  | Age (Yr's) | -0.0008 | 55473 | 0.302 | 1 | -0.08 | 31 | 31 |
|  |  | Sex (F) | -0.0294 | 55473 | 0.262 | 1 | -0.15 | 31 | 31 |
|  |  | BMI (kg/m2) | -0.0002 | 55473 | 0.865 | 1 | -0.02 | 31 | 31 |
|  | Duration | Intercept | 1.2261 | 55473 | 0.257 | 1 | 0.30 | 55484 | 31 |
|  |  | Time into Trip | -0.0083 | 55473 | 0.001 | 0.009 | -0.04 | 55484 | 31 |
|  |  | Trip Duration | 0.0212 | 55473 | 3.19E-22 | 2.87E-21 | 0.11 | 2231 | 31 |
|  |  | CPAP use (2 night mean) | 0.0007 | 55473 | 1.47E-05 | 0.0001 | 0.05 | 612 | 31 |
|  |  | Days on CPAP | -0.0059 | 55473 | 1.43E-05 | 0.0001 | -0.05 | 612 | 31 |
|  |  | Baseline mean | 0.7168 | 55473 | 1.32E-15 | 1.18E-14 | 0.18 | 31 | 31 |
|  |  | ODI | 7.80E-05 | 55473 | 0.988 | 1 | 0.001 | 31 | 31 |
|  |  | ESS | -0.0156 | 55473 | 0.370 | 1 | -0.04 | 31 | 31 |
|  |  | Age (Yr's) | 0.0075 | 55473 | 0.359 | 1 | 0.03 | 31 | 31 |
|  |  | Sex (F) | -0.3824 | 55473 | 0.142 | 1 | -0.09 | 31 | 31 |
|  |  | BMI (kg/m2) | 0.0077 | 55473 | 0.572 | 1 | 0.04 | 31 | 31 |
|  | Sig Frequency | Intercept | -0.1036 | 2221 | 0.740 | 1 | -0.28 | 2231 | 31 |
|  |  | Trip Duration | -0.0066 | 2221 | 6.29E-14 | 5.66E-13 | -0.34 | 2231 | 31 |
|  |  | CPAP use (2 night mean) | 8.99E-05 | 2221 | 0.225 | 1 | 0.07 | 612 | 31 |
|  |  | Days on CPAP | -0.0010 | 2221 | 0.095 | 0.854 | -0.10 | 612 | 31 |
|  |  | Baseline mean | 0.8740 | 2221 | 6.40E-10 | 5.76E-09 | 2.33 | 31 | 31 |
|  |  | ODI | 0.0062 | 2221 | 0.005 | 0.047 | 0.44 | 31 | 31 |
|  |  | ESS | 0.0068 | 2221 | 0.329 | 1 | 0.20 | 31 | 31 |
|  |  | Age (Yr's) | -0.0011 | 2221 | 0.735 | 1 | -0.06 | 31 | 31 |
|  |  | Sex (F) | -0.0656 | 2221 | 0.528 | 1 | -0.17 | 31 | 31 |
|  |  | BMI (kg/m2) | 0.0008 | 2221 | 0.888 | 1 | 0.04 | 31 | 31 |

**Supplementary Table f:** CPAP Compliance Results (Previous 3 night’s mean usage)

| Event | Feature | Predictor | Estimate | DF | P | Adjusted P | Effect Size (LME) | Data Points | Pt (n) |
| --- | --- | --- | --- | --- | --- | --- | --- | --- | --- |
| Brake | Magnitude | Intercept | -0.0285 | 25200 | 0.713 | 1 | -0.23 | 25211 | 31 |
|  |  | Time into Trip | -0.0005 | 25200 | 5.69E-06 | 5.12E-05 | -0.07 | 25211 | 31 |
|  |  | Trip Duration | 0.0002 | 25200 | 0.097 | 0.874 | 0.03 | 2230 | 31 |
|  |  | CPAP use (3 night mean) | -2.83E-05 | 25200 | 0.0002 | 0.002 | -0.07 | 612 | 31 |
|  |  | Days on CPAP | 0.0001 | 25200 | 0.025 | 0.222 | 0.04 | 612 | 31 |
|  |  | Baseline mean | 0.9821 | 25200 | 1.05E-06 | 9.46E-06 | 8.09 | 31 | 31 |
|  |  | ODI | 0.0008 | 25200 | 0.003 | 0.028 | 0.18 | 31 | 31 |
|  |  | ESS | -0.0004 | 25200 | 0.648 | 1 | -0.04 | 31 | 31 |
|  |  | Age (Yr's) | 0.0002 | 25200 | 0.551 | 1 | 0.04 | 31 | 31 |
|  |  | Sex (F) | 0.0003 | 25200 | 0.980 | 1 | 0.003 | 31 | 31 |
|  |  | BMI (kg/m2) | 0.0003 | 25200 | 0.626 | 1 | 0.05 | 31 | 31 |
|  | Duration | Intercept | 0.7287 | 25200 | 0.115 | 1 | 0.20 | 25211 | 31 |
|  |  | Time into Trip | -0.0233 | 25200 | 2.96E-12 | 2.66E-11 | -0.11 | 25211 | 31 |
|  |  | Trip Duration | 0.0337 | 25200 | 3.16E-32 | 2.85E-31 | 0.19 | 2230 | 31 |
|  |  | CPAP use (3 night mean) | 0.0003 | 25200 | 0.110 | 0.988 | 0.02 | 612 | 31 |
|  |  | Days on CPAP | -0.0066 | 25200 | 6.23E-06 | 5.61E-05 | -0.07 | 612 | 31 |
|  |  | Baseline mean | 0.8122 | 25200 | 2.38E-61 | 2.14E-60 | 0.23 | 31 | 31 |
|  |  | ODI | 0.0058 | 25200 | 0.003 | 0.023 | 0.04 | 31 | 31 |
|  |  | ESS | -0.0136 | 25200 | 0.010 | 0.088 | -0.04 | 31 | 31 |
|  |  | Age (Yr's) | 0.0046 | 25200 | 0.169 | 1 | 0.02 | 31 | 31 |
|  |  | Sex (F) | 0.2190 | 25200 | 0.014 | 0.122 | 0.06 | 31 | 31 |
|  |  | BMI (kg/m2) | -0.0058 | 25200 | 0.208 | 1 | -0.03 | 31 | 31 |
|  | Sig Frequency | Intercept | 0.0612 | 2220 | 0.766 | 1 | 0.21 | 2230 | 31 |
|  |  | Trip Duration | -0.0067 | 2220 | 2.60E-22 | 2.34E-21 | -0.44 | 2230 | 31 |
|  |  | CPAP use (3 night mean) | -0.0002 | 2220 | 0.004 | 0.035 | -0.17 | 612 | 31 |
|  |  | Days on CPAP | 0.0011 | 2220 | 0.027 | 0.243 | 0.13 | 612 | 31 |
|  |  | Baseline mean | 0.8773 | 2220 | 2.71E-11 | 2.44E-10 | 3.02 | 31 | 31 |
|  |  | ODI | 0.0032 | 2220 | 0.031 | 0.281 | 0.29 | 31 | 31 |
|  |  | ESS | 0.0041 | 2220 | 0.378 | 1 | 0.16 | 31 | 31 |
|  |  | Age (Yr's) | -0.0016 | 2220 | 0.494 | 1 | -0.10 | 31 | 31 |
|  |  | Sex (F) | -0.0389 | 2220 | 0.582 | 1 | -0.13 | 31 | 31 |
|  |  | BMI (kg/m2) | 0.0014 | 2220 | 0.703 | 1 | 0.09 | 31 | 31 |
| Acceleration | Magnitude | Intercept | 0.0223 | 57364 | 0.735 | 1 | 0.19 | 57375 | 31 |
|  |  | Time into Trip | 0.0004 | 57364 | 9.94E-07 | 8.95E-06 | 0.05 | 57375 | 31 |
|  |  | Trip Duration | -0.0009 | 57364 | 5.24E-45 | 4.72E-44 | -0.16 | 2230 | 31 |
|  |  | CPAP use (3 night mean) | -1.76E-05 | 57364 | 0.000 | 0.004 | -0.04 | 612 | 31 |
|  |  | Days on CPAP | 9.75E-05 | 57364 | 0.011 | 0.103 | 0.03 | 612 | 31 |
|  |  | Baseline mean | 0.8137 | 57364 | 0.001 | 0.008 | 6.93 | 31 | 31 |
|  |  | ODI | 0.0006 | 57364 | 0.050 | 0.446 | 0.15 | 31 | 31 |
|  |  | ESS | 0.0007 | 57364 | 0.497 | 1 | 0.07 | 31 | 31 |
|  |  | Age (Yr's) | -1.45E-05 | 57364 | 0.976 | 1 | -0.002 | 31 | 31 |
|  |  | Sex (F) | -0.0174 | 57364 | 0.291 | 1 | -0.15 | 31 | 31 |
|  |  | BMI (kg/m2) | 0.0002 | 57364 | 0.854 | 1 | 0.03 | 31 | 31 |
|  | Duration | Intercept | 0.6529 | 57364 | 0.001 | 0.013 | 0.19 | 57375 | 31 |
|  |  | Time into Trip | -0.0154 | 57364 | 1.75E-13 | 1.57E-12 | -0.08 | 57375 | 31 |
|  |  | Trip Duration | 0.0100 | 57364 | 1.43E-08 | 1.28E-07 | 0.06 | 2230 | 31 |
|  |  | CPAP use (3 night mean) | -6.47E-05 | 57364 | 0.581 | 1 | -0.01 | 612 | 31 |
|  |  | Days on CPAP | 0.0019 | 57364 | 0.042 | 0.382 | 0.02 | 612 | 31 |
|  |  | Baseline mean | 0.8317 | 57364 | 3.1E-262 | 2.77E-261 | 0.24 | 31 | 31 |
|  |  | ODI | -0.0002 | 57364 | 0.877 | 1 | -0.001 | 31 | 31 |
|  |  | ESS | 0.0201 | 57364 | 3.50E-09 | 3.15E-08 | 0.06 | 31 | 31 |
|  |  | Age (Yr's) | -0.0031 | 57364 | 0.134 | 1 | -0.02 | 31 | 31 |
|  |  | Sex (F) | 0.1955 | 57364 | 0.0003 | 0.002 | 0.06 | 31 | 31 |
|  |  | BMI (kg/m2) | -0.0013 | 57364 | 0.668 | 1 | -0.01 | 31 | 31 |
|  | Sig Frequency | Intercept | -0.0592 | 2220 | 0.862 | 1 | -0.16 | 2230 | 31 |
|  |  | Trip Duration | -0.0086 | 2220 | 1.64E-21 | 1.48E-20 | -0.43 | 2230 | 31 |
|  |  | CPAP use (3 night mean) | -0.0001 | 2220 | 0.160 | 1 | -0.08 | 612 | 31 |
|  |  | Days on CPAP | 0.0005 | 2220 | 0.385 | 1 | 0.05 | 612 | 31 |
|  |  | Baseline mean | 0.7917 | 2220 | 4.60E-06 | 4.14E-05 | 2.08 | 31 | 31 |
|  |  | ODI | 0.0048 | 2220 | 0.041 | 0.367 | 0.34 | 31 | 31 |
|  |  | ESS | 0.0091 | 2220 | 0.245 | 1 | 0.26 | 31 | 31 |
|  |  | Age (Yr's) | 0.0005 | 2220 | 0.881 | 1 | 0.03 | 31 | 31 |
|  |  | Sex (F) | -0.0901 | 2220 | 0.453 | 1 | -0.24 | 31 | 31 |
|  |  | BMI (kg/m2) | 0.0004 | 2220 | 0.945 | 1 | 0.02 | 31 | 31 |
| Turn | Magnitude | Intercept | -0.0150 | 55473 | 0.878 | 1 | -0.07 | 55484 | 31 |
|  |  | Time into Trip | -8.17E-05 | 55473 | 0.508 | 1 | -0.01 | 55484 | 31 |
|  |  | Trip Duration | 5.06E-05 | 55473 | 0.638 | 1 | 0.01 | 2231 | 31 |
|  |  | CPAP use (3 night mean) | -2.01E-05 | 55473 | 0.027 | 0.239 | -0.03 | 612 | 31 |
|  |  | Days on CPAP | -0.0002 | 55473 | 0.003 | 0.027 | -0.04 | 612 | 31 |
|  |  | Baseline mean | 1.0016 | 55473 | 2.09E-06 | 1.88E-05 | 4.99 | 31 | 31 |
|  |  | ODI | 0.0017 | 55473 | 0.002 | 0.014 | 0.26 | 31 | 31 |
|  |  | ESS | 0.0019 | 55473 | 0.295 | 1 | 0.10 | 31 | 31 |
|  |  | Age (Yr's) | -0.0009 | 55473 | 0.284 | 1 | -0.08 | 31 | 31 |
|  |  | Sex (F) | -0.0288 | 55473 | 0.275 | 1 | -0.14 | 31 | 31 |
|  |  | BMI (kg/m2) | -0.0002 | 55473 | 0.908 | 1 | -0.02 | 31 | 31 |
|  | Duration | Intercept | 1.2940 | 55473 | 0.229 | 1 | 0.32 | 55484 | 31 |
|  |  | Time into Trip | -0.0083 | 55473 | 0.001 | 0.008 | -0.04 | 55484 | 31 |
|  |  | Trip Duration | 0.0212 | 55473 | 4.39E-22 | 3.95E-21 | 0.11 | 2231 | 31 |
|  |  | CPAP use (3 night mean) | 0.0007 | 55473 | 0.0002 | 0.002 | 0.05 | 612 | 31 |
|  |  | Days on CPAP | -0.0062 | 55473 | 6.49E-06 | 5.84E-05 | -0.05 | 612 | 31 |
|  |  | Baseline mean | 0.7146 | 55473 | 1.16E-15 | 1.05E-14 | 0.18 | 31 | 31 |
|  |  | ODI | 2.42E-05 | 55473 | 0.996 | 1 | 0.0002 | 31 | 31 |
|  |  | ESS | -0.0158 | 55473 | 0.361 | 1 | -0.04 | 31 | 31 |
|  |  | Age (Yr's) | 0.0071 | 55473 | 0.387 | 1 | 0.03 | 31 | 31 |
|  |  | Sex (F) | -0.3788 | 55473 | 0.144 | 1 | -0.09 | 31 | 31 |
|  |  | BMI (kg/m2) | 0.0078 | 55473 | 0.561 | 1 | 0.04 | 31 | 31 |
|  | Sig<br>Frequency | Intercept | -0.0892 | 2221 | 0.776 | 1 | -0.24 | 2231 | 31 |
|  |  | Trip Duration | -0.0066 | 2221 | 5.91E-14 | 5.31E-13 | -0.34 | 2231 | 31 |
|  |  | CPAP use (3 night mean) | 6.06E-05 | 2221 | 0.445 | 1 | 0.04 | 612 | 31 |
|  |  | Days on CPAP | -0.0010 | 2221 | 0.101 | 0.906 | -0.10 | 612 | 31 |
|  |  | Baseline mean | 0.8720 | 2221 | 8.05E-10 | 7.25E-09 | 2.32 | 31 | 31 |
|  |  | ODI | 0.0062 | 2221 | 0.005 | 0.049 | 0.44 | 31 | 31 |
|  |  | ESS | 0.0067 | 2221 | 0.342 | 1 | 0.20 | 31 | 31 |
|  |  | Age (Yr's) | -0.0012 | 2221 | 0.712 | 1 | -0.06 | 31 | 31 |
|  |  | Sex (F) | -0.0643 | 2221 | 0.537 | 1 | -0.17 | 31 | 31 |
|  |  | BMI (kg/m2) | 0.0009 | 2221 | 0.871 | 1 | 0.04 | 31 | 31 |

**Supplementary Table g:**
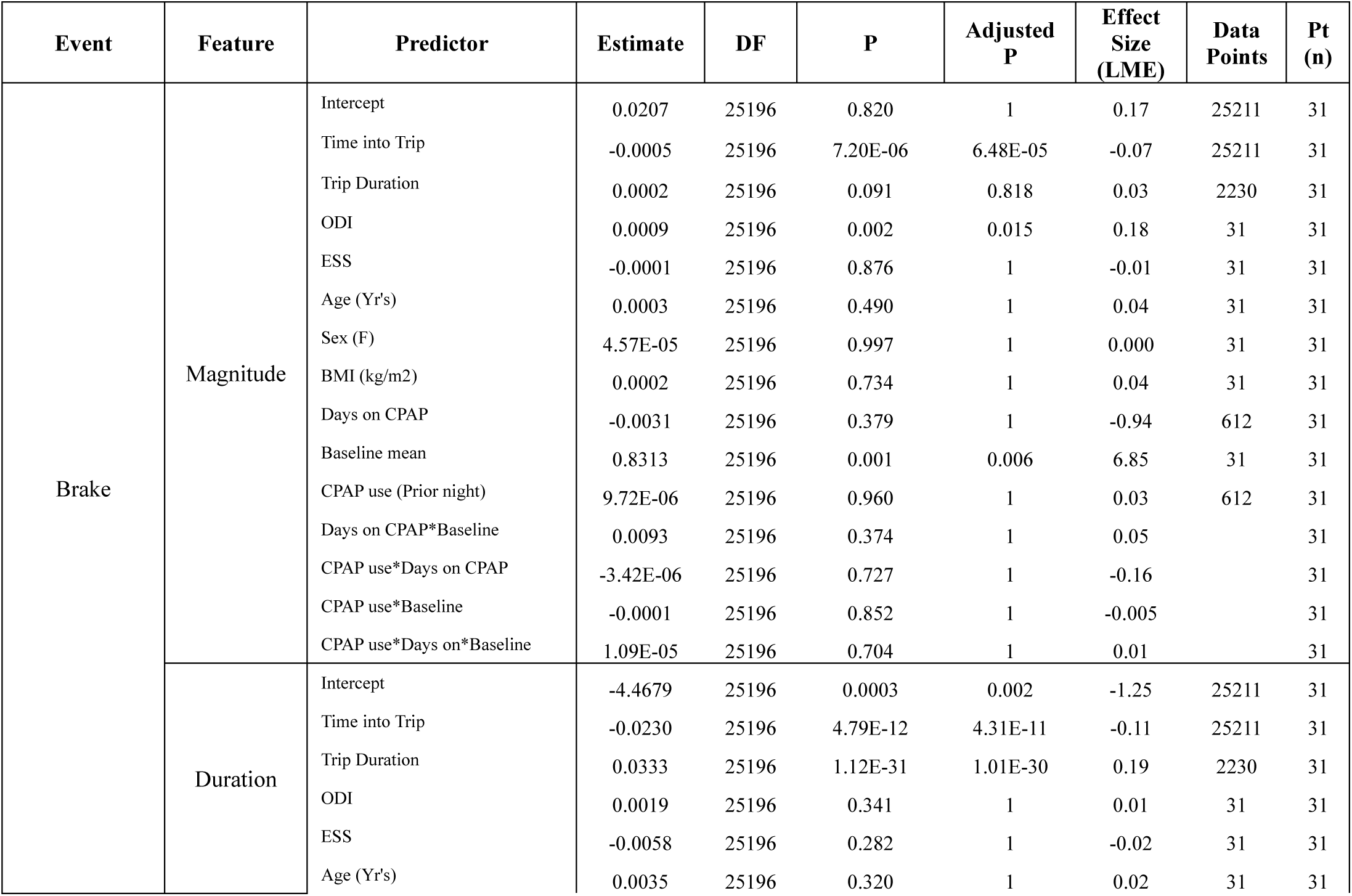
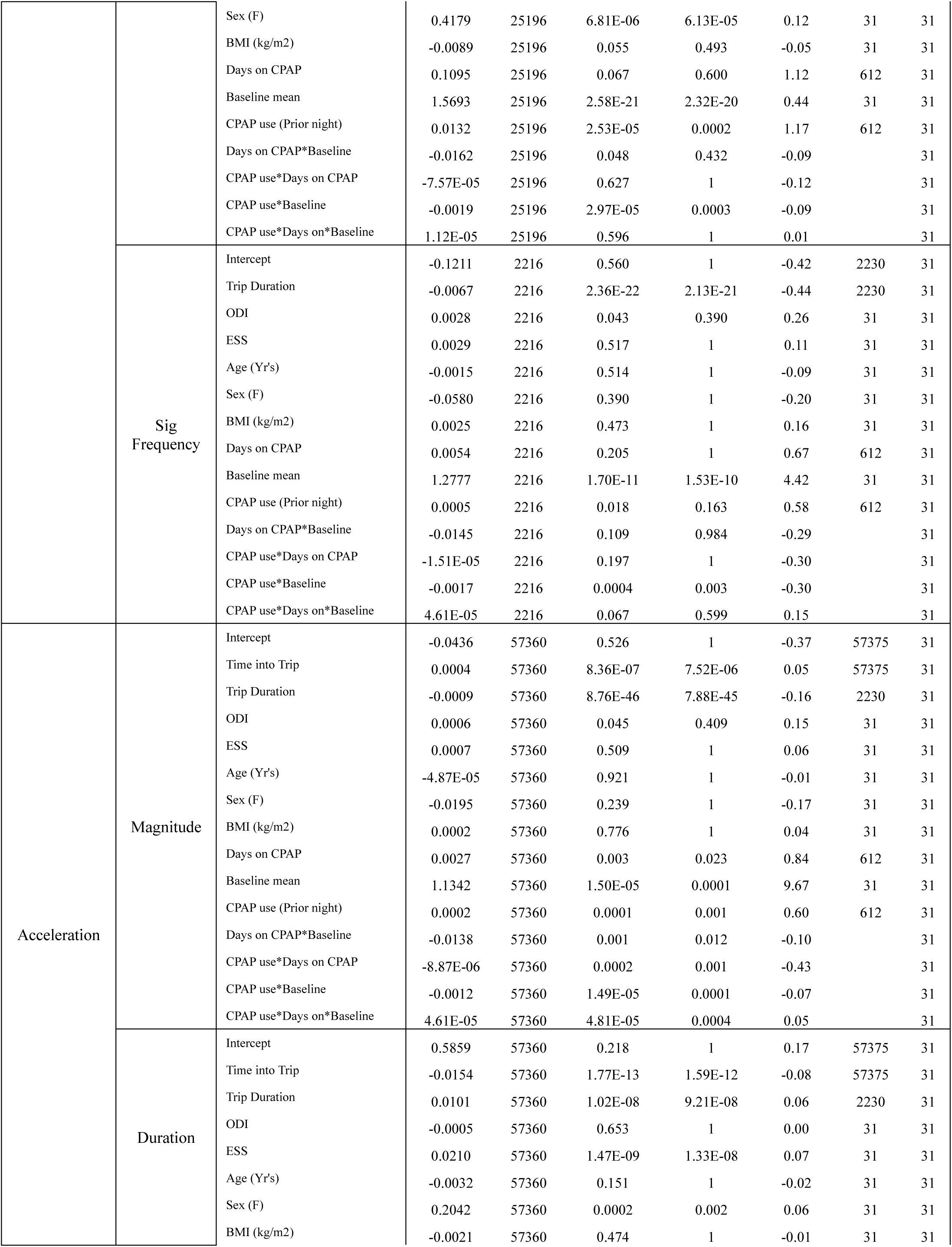
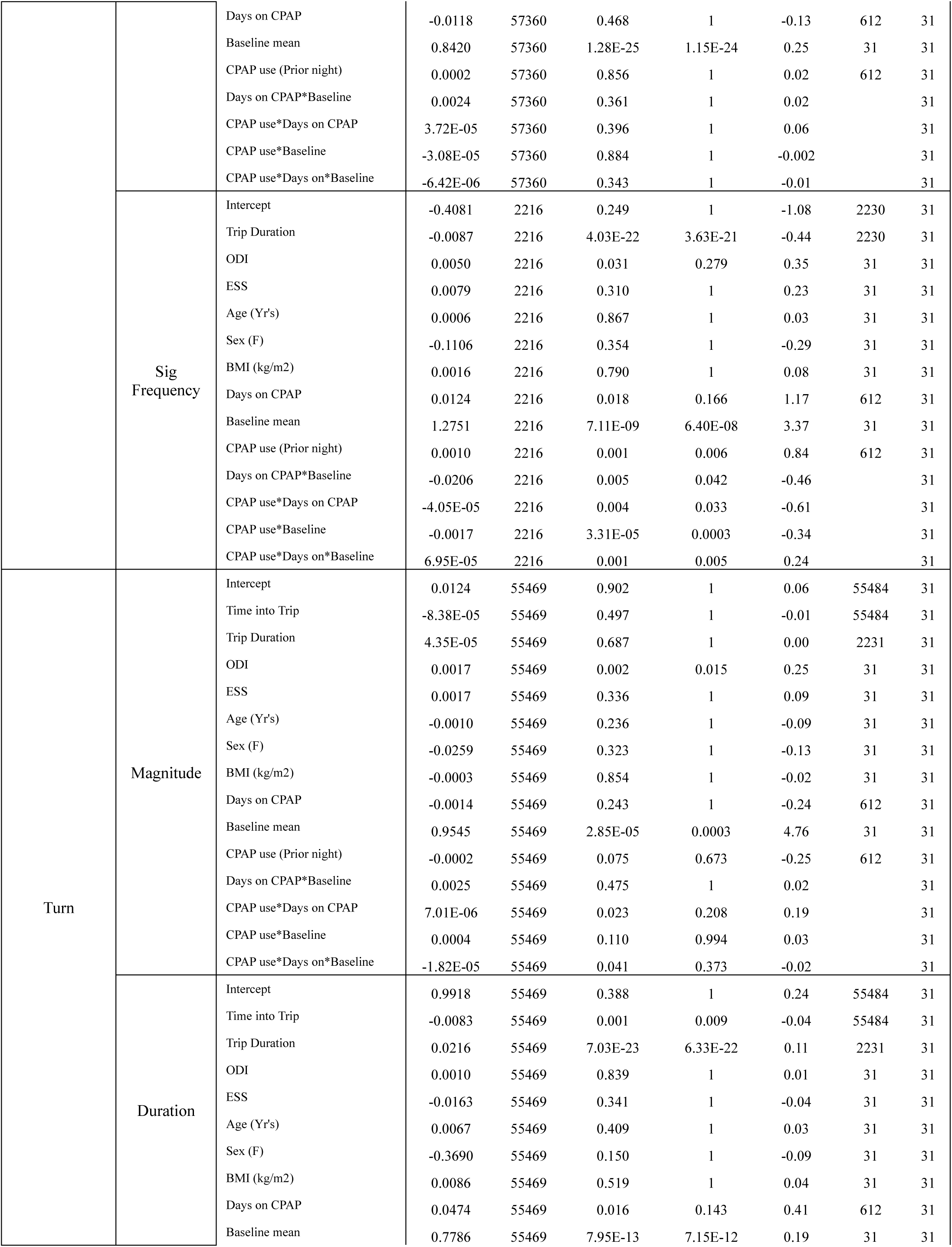
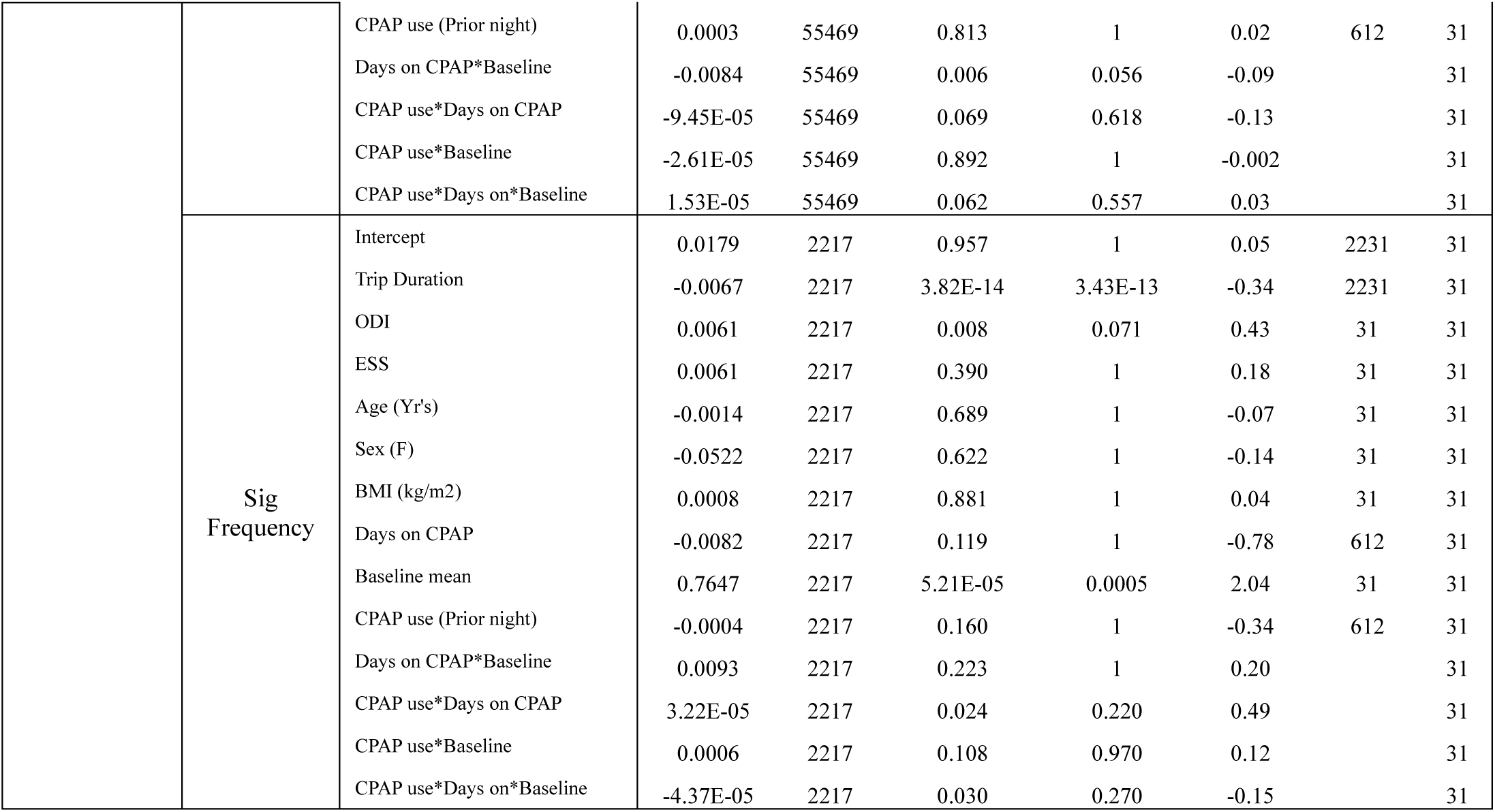
Baseline and CPAP use interaction LME Results (Prior night’s usage)

